# Present-day admixed genomes reveal prehistoric adaptation to cold, maritime diet, and local pathogens in Patagonia

**DOI:** 10.1101/2025.08.21.25334163

**Authors:** Patricio Pezo-Valderrama, Michael Orellana-Soto, Alexandra Salcedo, Juan Esteban Rodríguez-Rodríguez, Cristian Yañez, Elena Llop, Eugenio Aspillaga, Maria Laura Parolin, Andrés Moreno-Estrada, Carlos D. Bustamante, Julian R. Homburger, Christopher R. Gignoux, Alexander Ioannidis, Celeste Eng, Scott Huntsman, Esteban G. Burchard, Tábita Hünemeier, Mauricio Moraga, Ricardo A. Verdugo

## Abstract

Patagonia, the last region settled by humans after the out-of-Africa migration, provides a unique case for studying genetic adaptations to extreme environments. Despite archaeological evidence of human occupation for at least 13,500 years, the genetic history of Indigenous Patagonians remains understudied. Here, we analyze genome-wide data from individuals with high Native American ancestry to investigate their genetic structure, admixture history, and response to environmental pressures through selection. Our results reveal widespread European admixture, which in Chilean Patagonia started 5 generations earlier than in the capital of the colony, suggesting prolonged European contact relative to lower latitudes. We identified a strong north-south genomic differentiation, where common ancestry with Mapuche people is observed in northern Patagonia while Tierra del Fuego shows a unique component of genetic variation. We found selection signals predating European contact, possibly shaped during the settlement of native ancestors in Patagonia. These signals support adaptation to extreme cold and maritime diet, including genomic regions associated with height (*SUPT3H, CADM1, RUNX2*), lipid metabolism (*MSMO1, SORT1/PSCR2, CELSR2*), and energy homeostasis (*ZNF202*). Furthermore, enrichment of Native American ancestry was observed in the MHC complex, likely representing the genetic footprint of past selection events driven by adaptation to endemic pathogens that were still beneficial after admixture with Europeans. These findings provide novel insights into the genetic legacy of Patagonian populations, demonstrating how natural selection has shaped their resilience to environmental challenges and the dynamics of demographic changes over time.

## Introduction

Patagonia, the southernmost region of human dispersal following the out-of-Africa migration, provides critical insights into the peopling of the Americas. Archaeological evidence indicates initial human occupation around 13,500 years before present (YBP) (Borrero et al., 1998). Situated south of 40°S latitude in South America, this region spans from the Pacific to the Atlantic Ocean and includes Tierra del Fuego, characterized by extreme environmental conditions and diverse geography (Garreaud et al., 2013). The native inhabitants of Patagonia demonstrated remarkable resilience, adapting to harsh climates and significant dietary pressures (Rivas et al., 1999; Yesner et al., 2003; Orquera & Piana, 2009; Orquera et al., 2011; Báez et al., 2015). However, reconstructing the genetic history of Native American populations in Patagonia is challenging due to their widespread population decline following European contact, cultural discontinuity, and extensive admixture (de la Fuente et al., 2018; Parolin et al., 2019).

As humans migrated from Beringia to Tierra del Fuego, they encountered diverse environmental pressures, including high altitudes, novel disease vectors, unique dietary resources, and intense UV radiation in subpolar latitudes, all of which likely drove biological adaptations (Lindo and DeGiorgio, 2021). Genomic studies of Native American populations have identified signatures of selection associated with adaptations to cold climates and marine diets, particularly in genes such as *THADA*, *PRKG1*, *CPT1A*, and *FADS1/FADS2* (Cardona et al., 2014; Clemente et al., 2014; Fumagalli et al., 2015). Notably, alleles in the FADS gene cluster, which play a crucial role in lipid metabolism, exhibit higher frequencies in lower-latitude Native populations, reflecting past selective pressures for cold adaptation (Amorim et al., 2017). These genetic variants also influence lipid regulation in admixed populations with Native ancestry (K. M. Reynolds et al., 2023).

Genetic evidence of local adaptations has also been documented in Peruvian populations. High-altitude Andean groups exhibit selection signals in hypoxia-related genes, including *EGLN1*, *ENDRA*, *PRKAA1*, *NOS2A, TGFA, APIP,* and *FAM213A* among Quechua, Aymara, and admixed populations from Puno, Lima, and Cusco (Bigham et al., 2009, 2010; Valverde et al., 2015; Caro-Consuegra et al., 2022). Selection for arsenic metabolism has also been detected in the *ASM3MT* gene among Andean populations (Schlebusch et al., 2013, 2015), while adaptation related to cardiovascular function (*HAND2-AS1*, *TLL1*, *DUSP27*, *TBX5*, *PLXNA4*, and *SGCD*), skin pigmentation (*MITF*), and glucose metabolism (*GLIS3*, *PPP1R3C*, and *GANC*) have been reported (Borda et al., 2020; Caro-Consuegra et al., 2022). Coastal and rainforest Peruvian populations exhibit immune-related adaptations (*SIGLEC8*, *TRIM21*, *CD44*, *ICAM*, *CBLB*, *PRDM1*, *BRD2*, and *HLA* genes), likely driven by pathogen exposure (Caro-Consuegra et al., 2022). Similarly, Amazonian populations exhibit selection signals in genes associated with antigen recognition and resistance to *Trypanosoma cruzi*, the causative agent of Chagas disease (Borda et al., 2020; Couto-Silva et al., 2023). Post-admixture selection has been observed to increase the frequency of specific ancestries at certain loci, particularly in immune-related genes such as those in the Major Histocompatibility Complex (MHC) (Jeong & Di Rienzo, 2014; Lindo et al., 2016; Vicuña et al., 2020; Q. Zhou et al., 2016). In the Chilean admixed population, selection signals have been detected in genes associated with pigmentation, thermogenesis, and immune response among Mapuche populations (Vicuña et al., 2020), as well as arsenic metabolism adaptations in Andean populations from the Atacama Desert (Vicuña et al., 2019). These findings underscore the significance of genomic studies in elucidating both ancient and recent evolutionary processes, as well as their influence on the health of contemporary populations. However, no studies have yet explored adaptive selection in Indigenous populations of Patagonia and Tierra del Fuego.

The origin of Patagonia’s first inhabitants remains debated, with hypotheses suggesting migration via the Pacific coast or from highland Andean populations (Rothhammer & Dillehay, 2009; de la Fuente et al., 2018; Arango-Isaza et al., 2023). By 7,300 cal BP, western Patagonia saw a transition to a maritime lifestyle, evidenced by early settlements in the Magallanes Strait (Punta Santa Ana) and south of Beagle Channel (Imiwa I and Lancha Packewaia) (Legoupil & Fontugne, 1997). Alternative hypotheses propose an independent origin of maritime populations in northwest Patagonia (Isla Grande de Chiloé and Seno de Reloncaví) around 6,500 cal BP (Rivas et al., 1999; Gaete et al., 2004; Ocampo & Rivas, 2004). Stable isotope analysis indicates that western Patagonia settlers relied primarily on marine resources, while eastern continental groups depended on terrestrial sources, such as guanaco, rodents, and birds (Yesner et al., 2003; Panarello et al., 2006; Tessone et al., 2009; Berón et al., 2009; Borrero et al., 2009; Porras et al., 2014; Báez et al., 2015).

During the colonial period (17^th^-19^th^ centuries), extensive admixture occurred between Indigenous populations, European settlers, and African forced migrants (Gravel et al., 2013; Homburger et al., 2015; Adhikari et al., 2017). In Chile, genomic studies reveal relatively equal contributions from European and Native American ancestry, although Native ancestry is higher in Chilean Patagonia (46%) compared to Argentine Patagonia (35.8%) (Eyheramendy et al., 2015; Verdugo et al., 2020). It is reasonable to hypothesize that if selection has occurred in Patagonia after admixture, where inherited haplotypes from either ancestry would provide an adaptive advantage, it would leave a signature of enrichment of that ancestry in the loci under selection.

In this study, we investigate genomic evidence for genetic adaptations in populations native to Patagonia. We analyzed genome-wide data from individuals with high Native American Ancestry, including 50 newly genotyped individuals and 24 previously reported (de la Fuente et al., 2018). Our findings provide novel insights into pre- and post-admixture selection in Patagonia, shedding light on the genetic adaptations in the last region colonized by humans.

## Results

### Patagonian Populations

Seven locations across Patagonia were sampled, encompassing both high- and low-latitude regions in Chile and Argentina (Fig. 1). Genotyping was conducted on 50 individuals with Native American ancestry, previously recruited from Laitec island in the Chiloé Archipelago (n=30), and four locations in Argentinean Patagonia (n=20). Genotyping was performed using the Axiom Latino Array (World Array 4, Affymetrix Inc., Santa Clara, CA), yielding genotypes for 607,853 single nucleotide polymorphisms (SNPs) that passed quality control filtering. Two individuals with Chilean and Bolivian parental ancestry and one individual with > 5% of missingness were excluded from the Argentinian dataset, resulting in a final dataset of 47 individuals. Additionally, genotype data from 24 individuals from Punta Arenas (Kawésqar) and Villa Ukika (Yámana), previously analyzed in de la Fuente Castro et al., (2018), were incorporated. These latter individuals inhabit ancestral territories historically associated with maritime hunter-gatherer populations (Martinic, 2004). Self-reported ancestry indicated descent from the Kawésqar and Yámana populations, respectively. In contrast, individuals from Laitec, a rural locality, did not self-identify with a specific Indigenous group; however, genetic and ethnographic evidence suggests a potential ancestral connection to the extinct Chono population of northern Patagonia (García et al., 2006).

**Figure 1.**
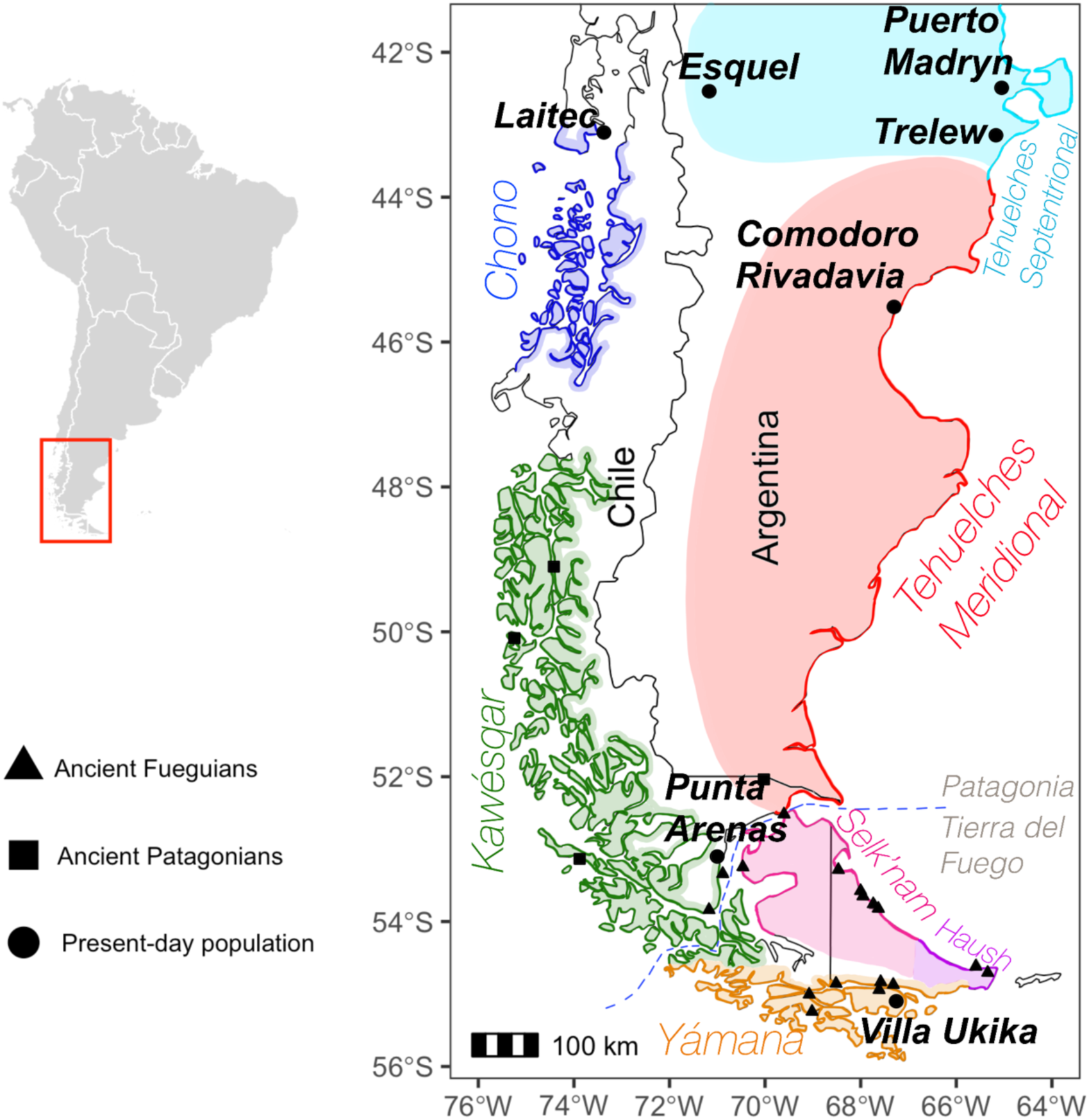
Patagonia geographic area in South America. Sampling location for individuals with self-reported and putative Native ancestry who were genotyped in this study are indicated (black dots). Esquel, Puerto Madryn, Trelew, and Comodoro Rivadavia were grouped as Argentinian Patagonia. Geo-localization of archeological site in Chilean Patagonia (square) and Tierra del Fuego (triangles) for published ancient DNA data is also indicated (Raghavan et al., 2015; Moreno-Mayar et al., 2018; de la Fuente et al., 2018; Nakatsuka et al., 2020). The historical dispersal range for Patagonian native populations was described according to ethnographic evidence (Martinic, 2004; Casamiquela, 2007).

### Genetic structure and ancestral components in Patagonia

To investigate population structure, we conducted Principal Component Analysis (PCA) and *ADMIXTURE* analyses on 74 unrelated individuals from contemporary Patagonian populations, alongside 20 other South American populations, 36 ancient DNA samples from Patagonia, and references individuals from the 1000 Genomes Project (*see Supplementary Note 6, Figure S1*). These analyses revealed a genetic gradient between European and Native American ancestry, with the latter predominating (*see Supplementary Note 7, Figure S2*). Given the negligible African and Asian contributions, we excluded these reference populations and re-ran the PCA. The first two principal components distinguished four major clusters corresponding to geographic regions in South America: (1) Andean (Quechua, Puno, Aymara, and Diaguita); (2) Central-South Chile and Patagonian (Pehuenche, Huilliche, Laitec, Kawesqar and Argentinean Patagonians from Esquel, Puerto Madryn, Trelew, and Comodoro Rivadavia); (3) other native groups from lowland South America (Arhuaco, Kogi, Wayuu, Embera, Guahibo, Waunana, Palikur, Piapoco, Inga, Ticuna, Chane, Wichi, Guarani, and Toba); (4) and Tierra del Fuego (Yamana, ancient Patagonians, and ancient Fueguians) (Fig. 2A).

**Figure 2.**
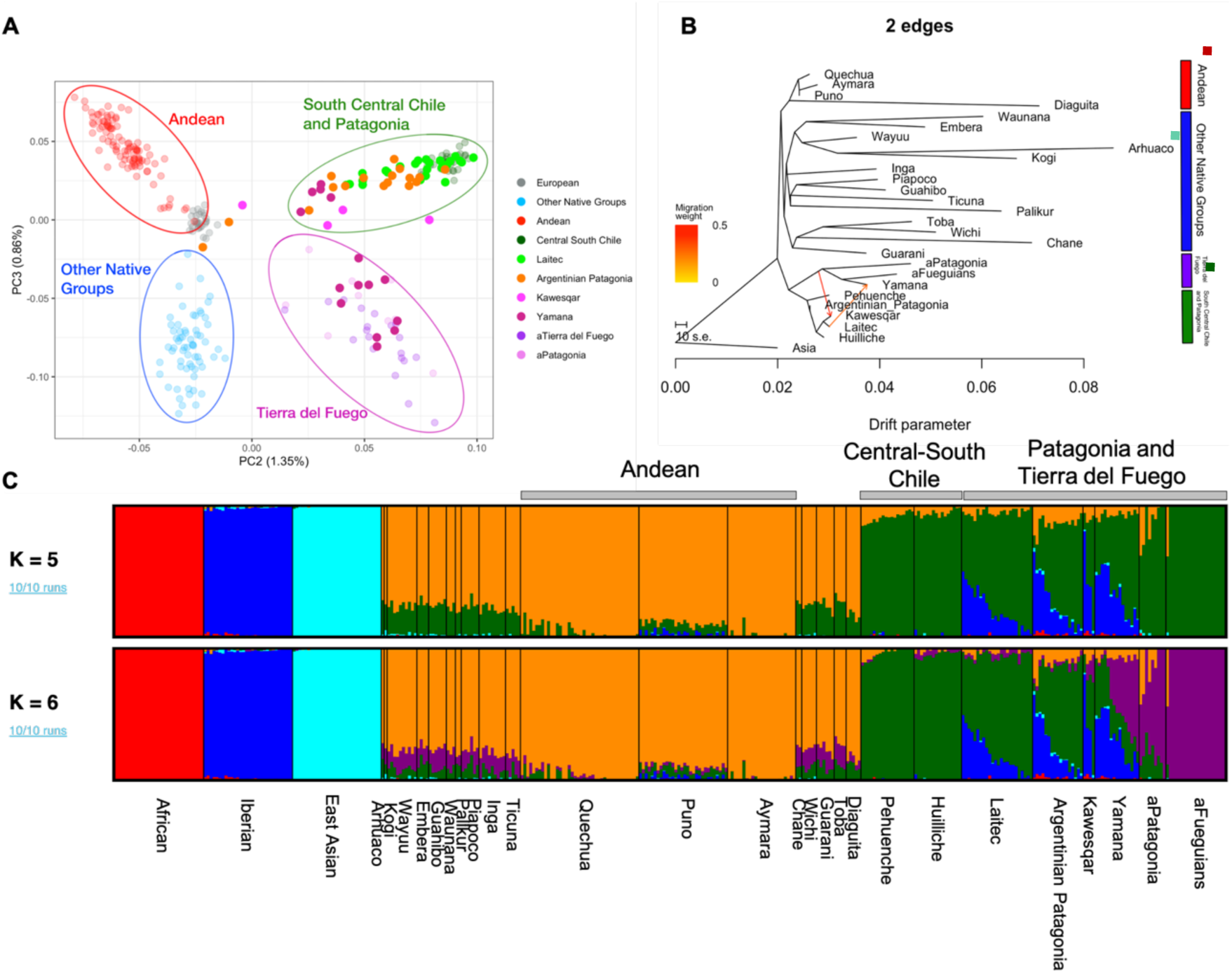
Population Structure in Patagonian samples. A) Principal Component Analysis (PCA) for 20 populations from South America (Reich et al., 2012), 36 ancient DNA samples from Patagonia (Raghavan et al., 2015; de la Fuente et al., 2018; Moreno-Mayar et al., 2018; Nakatsuka et al., 2020), and the European (IBS) reference population from 1000G (Auton & Salcedo, 2015). Each dot represents an individual from the following groups: Andean (Quechua, Puno, Aymara, and Diaguita), Central-South Chile (Pehuenche and Huilliche), other native groups from lowland South America (Arhuaco, Kogi, Wayuu, Embera, Guahibo, Waunana, Palikur, Piapoco, Inga, Ticuna, Chane, Wichi, Guarani, and Toba), Chilean Patagonians (Laitec, Kawésqar, Yamana), Argentinian Patagonians (Esquel, Puerto Madryn, Trelew, and Comodoro Rivadavia) and ancient samples from Patagonia (aPatagonian) and Tierra del Fuego (aFueguian). B), inferred maximum likelihood tree of 29 populations with two migration edges through *Treemix v1.13*. Migration arrows are colored according to their weight, and horizontal branch lengths are proportional to the amount of genetic drift that has occurred on the branch. The scale bar shows ten times the average standard error of the entries in the sample covariance matrix (W). Residual tree and likelihood value distribution are in *Supplementary Figures S5-6*. C). Each cluster is groups into Andean (red bar), Other Native Groups (blue bar), South Central Chile and Patagonia (green bar), and Tierra del Fuego (purple bar) Global ancestries by *ADMIXTURE*. Individuals were grouped by population (bottom) and sorted by latitude (north to south). Gray bars on top group samples by geographic location.

The *ADMIXTURE* analysis at *K=5*, which exhibited the lowest cross-validation error (*Supplementary Figure S3*), identified an ancestry composition primarily composed of Native American (74.2%) and European (24.3%) component, with minimal African admixture (<2%). A shared Native American ancestry component was prominent in all Patagonian populations, reaching its highest proportions in Central-South Chileans (94.4%, marked in green at K=5), and Argentinean Patagonia (61.1%), but declining in Chilean Patagonians of Kawésqar (41.3%) and Yámana (26.7%) descent (Fig. 2C). At *K=6*, an additional “Austral” ancestry component emerged (shown in purple), predominant in ancient Patagonian and Tierra del Fuego samples and highly frequent in modern Yámana (34.5%), but less so in Kawésqar (11.3%), Argentinian Patagonian (3.3%), and Mapuche (3.1%). Additionally, an Andean ancestry component was detected, with higher proportions observed in Argentinean Patagonia (10.4%), Kawésqar (6.7%), and Yámana (6.3%) compared to Laitec (1.7%) (Fig. 2C, *Supplementary Table S4, Figure S4*).

To assess genetic exchange among Patagonian populations, we applied a maximum-likelihood tree model using *Treemix v.1.13* (Pickrell & Pritchard, 2012), after masking non-Indigenous ancestry. This excluded 46.13% of the genome for the entire sample (*see Supplementary Note 5*). The extent of masking varied among groups, with the Kawésqar and Yámana populations having 57.59% and 51.30% of their genomes masked, respectively, while Laitec and Argentinean Patagonia groups exhibited masking levels of 35.12% and 40.53%, respectively.

The Argentinean Patagonia, Laitec, and Kawésqar populations clustered with Pehuenche and Huilliche, both part of the broader Mapuche cultural group in southern Chile and Argentina. In contrast, the Yámana clustered with ancient Patagonian and Tierra del Fuego individuals. The best-supported likelihood model (*Supplementary Figure S5-6*), identified two directional gene flow events: one from Tierra del Fuego cluster (ancient Patagonia, ancient Tierra del Fuego, and Yámana) into the Kawésqar population, and another from Laitec into the Yamana (Fig. 2B).

Local ancestry inference, performed using *RFMix v2* (Maples et al., 2013), indicated and average of 75.7% (SD = 5.2%) Native American and 23.8% (SD = 5.3%) European ancestry across all Patagonian individuals, consistent with ADMIXTURE estimates (Supplementary Tables S4, S5). To infer the timing of admixture events and migration patterns, we employed *TRACTs* (Gravel, 2012) to evaluate multiple demographic models (*see Supplementary Note 8, Figure S7-8, Table S6*). The results revealed distinct admixture histories among Patagonia populations. In Argentinean Patagonia and the Kawésqar, the best-fitting model suggested a single admixture event between Europeans and Native Americans approximately 9 and 10 generations ago, respectively, with no evidence of subsequent gene flow, indicating a stable genetic composition over time. In contrast, Laitec and Yámana exhibited older admixtures, dating to approximately 19 and 15 generations ago, respectively. Subsequent migratory pulses were detected, with Laitec incorporating African and additional Native American ancestry 17, and 7 generations ago, respectively, and Yámana receiving similar contributions 7, and 4 generations ago.

### Local ancestry enrichment

We assessed deviations from the genome-wide mean ancestry at each locus, identifying a locus within the major histocompatibility complex (MHC) haplotype spanning 3.05Mb (chr6:30.89-33.95). This region exhibited an exceptionally high frequency of Native American ancestry (98%; p-value 3,91E^-11^) across all Patagonian individuals (Fig. 3, Table S7). Additionally, we detected smaller peaks of Native American ancestry with significant deviation on chromosome 2, 12 and 21, each encompassing a restricted set of genes (Supplementary Table S7). In contrast, no significant selection signals were observed for European ancestry (mean: 23.8%). While substantial deviations from the genome-wide mean were noted for African ancestry (e.g., chromosome 15), this component was present at an extremely low proportion (mean: 0.4%) and was therefore excluded from further consideration.

**Figure 3.**
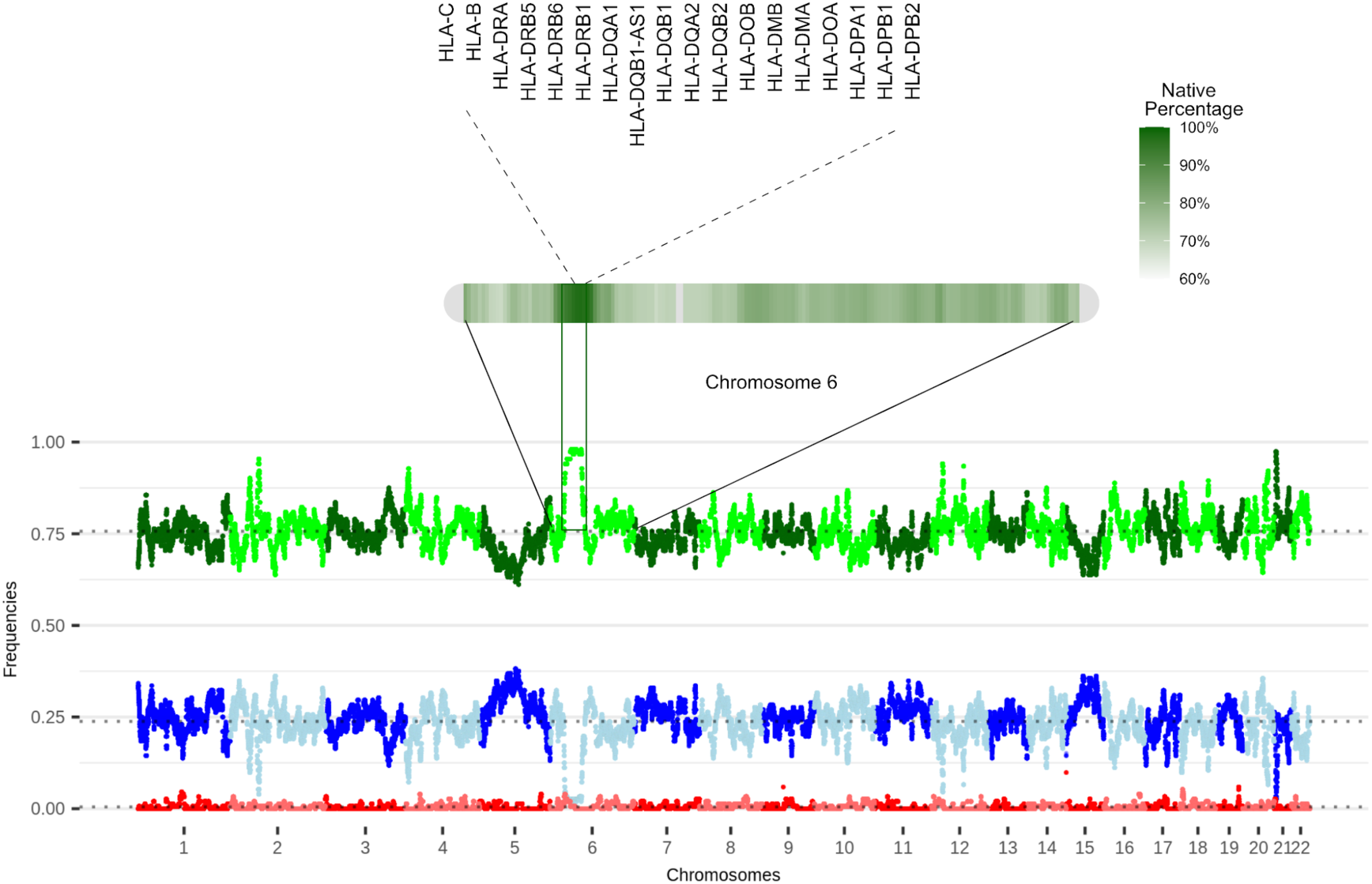
Local Ancestry Deviation along the genome for Patagonian Hunter-Gatherers descendants. In A), we show the Native American density along chromosome 6. Gene lists are displayed for the region with the highest proportion. For aesthetic purposes, we only incorporate the HLA cluster within this region. In B), the genome-ancestry proportions for Native American (green), European (blue), and African (red) ancestry are shown. The black dotted line represents the genome-wide average proportion for each ancestry.

### Signatures of positive selection in Native Patagonian genomes

We screened the Native American genomic portion of Chilean and Argentinean Patagonians populations (Fig. 4A-B) to identify genomic regions exhibiting signatures of selection. Using the normalized Population Branch Statistic (*PBSn1*) method (Yi et al., 2010; Crawford et al., 2017), we assessed allele frequency differentiation, while extended haplotypes were analyzed via the iHS statistic (Szpiech & Hernandez, 2014). Non-Indigenous chromosomal segments were masked, and individuals were categorized into two groups: (1) Patagonian Hunter-Gatherers’ descendants (PHG), which included Chilean Patagonians (Kawésqar and Yámana) and Argentinean Patagonians (Esquel, Puerto Madryn, Trelew, and Comodoro Rivadavia); and (2) Patagonian Maritime Hunter-Gatherers’ descendants (MHG), comprising only Chilean Patagonians. The top 1% of selection peaks were identified by comparing allele frequencies with African, European, and Native American reference panels. Functional annotation was performed using *Annovar* (Wang et al., 2010), with details in Supplementary Tables S8, S10, S12.

**Figure 4.**
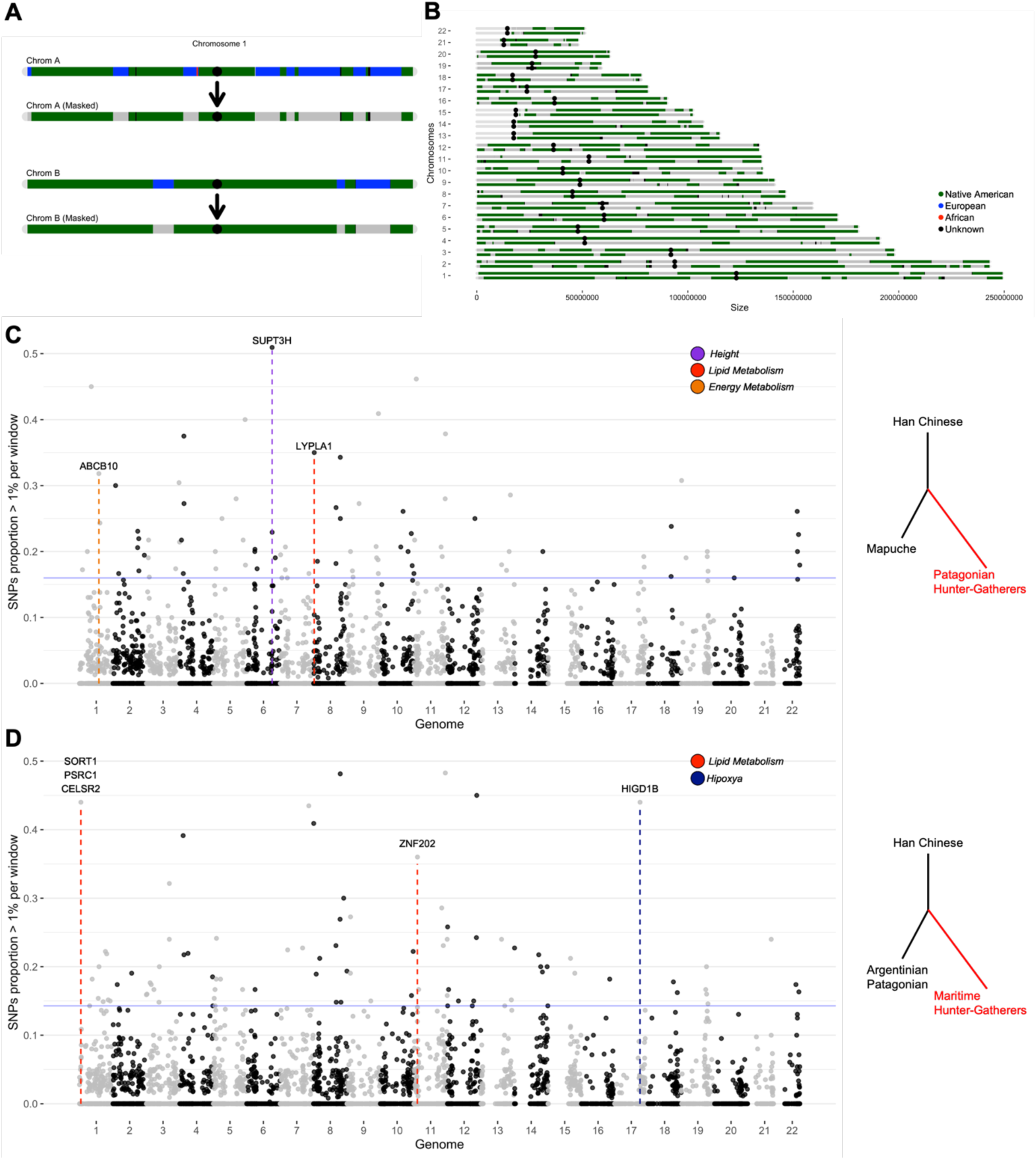
Genomic screens for signatures of positive selection in Native Patagonian genomes. A) Genome masking process is schematized in A and B, where all European (blue) and African (red) haplotypes are masked as missing data. B) An individual’s Native American genome after masking. Proportion of SNPs with a PBSn1 value within the top 1% genome-wide is shown for 200K-windows in C) Patagonian Hunter-Gatherers descendants and D) Maritime Hunter-Gatherers descendants. Both comparisons used Han Chinese as an outgroup. Patagonian Hunter-Gatherers (PGH) were compared to Pehuenche and Huilliche, who are members of the Mapuche People (MAP) as a close genetic group (EAS:MAP:PHG), while for Maritime Hunter-Gatherers (MHG), we used Argentinian Patagonia (EAS:ARP:MHG). The horizontal blue line indicates the top 1% of the distribution data. Among the top 10 windows in each comparison, those containing genes in categories that were enriched at the functional analysis were annotated with gene symbols. Colored vertical lines under those windows denote the functional category.

#### Patagonian Hunter-Gatherers’ descendants

Among the most prominent selection peaks in PHG, a highly differentiated intronic SNP (rs2322292) was detected within *MSMO1* at 4q32.3, a gene implicated in lipid biosynthesis, energy metabolism, obesity, and dyslipidemia (He et al., 2014; Lu et al., 2008). A strong selection signal was also observed in a 200Kb region containing *SUPT3H* at 6p211, previously associated with height variation in African Pygmy and Korean populations (J.-J. Kim et al., 2010; Mendizabal et al., 2012). Furthermore, selection peaks were identified in genomic regions harboring *LYPLA1* (8q11.23) and *ABCB10* (1q42.13), both implicated in lipid homeostasis and energy metabolism (Supplementary Table S8-9) (Seguin et al., 2017; Yano, 2017; Martinez et al., 2020) (Fig. 4C). The iHS analysis revealed an intronic variant (rs10495562) within *ADAM17*, a gene linked to lipid metabolism (Junyent et al., 2010; Kobayashi et al., 2016; Suzuki et al., 2016). Notably, *ADAM17* has been associated with convergent adaptation in light skin pigmentation in Asian populations (Norton et al., 2006). Moreover, two strong selection signals were identified on chromosome 6 within the *SUPT3H* gene (Supplementary Table S10, S11), reinforcing findings from the PBSn1 analysis.

#### Patagonian Maritime Hunter-Gatherers’ descendants

Although no cold-adaptation or maritime diet-related SNPs were detected via PBSn1 (Supplementary Table S12), several candidate regions were identified (Supplementary Table S13). Notably, the genomic region encompassing *SORT1*, *PSCR1*, and *CELSR2* at 1p.13.3, which play key roles in LDL cholesterol regulation and lipoprotein metabolism, showed signatures of selection (Arvind et al., 2014; Meroufel et al., 2015; Y.-J. Zhou et al., 2015). Additionally, selection evidence was observed in a genomic region containing *HIGD1B* at 17.q21.31, associated with hypoxia response (Salazar et al., 2020), and in *ZNF202* at 11q24.1, a gene implicated in lipid metabolism and energy homeostasis (Schmitz et al., 2004; Stene et al., 2006), also showed evidence of selection using PBSn1 (Fig. 4D).

#### Ancient Selection Signals

PBSn1 was also estimated in a set of whole-genome sequences (WGS) previously published for 36 samples of ancient DNA from Patagonia and Tierra del Fuego (Supplementary Table S3). We initially investigated genomic regions implicated in cold and maritime diet metabolism among Arctic populations, specially targeting *THADA*, *FADS*, *CPT1A*, and *LRP5* (Cardona et al., 2014; Clemente et al., 2014; Fumagalli et al., 2015). However, no extreme PBSn1 values were detected in 200kb-windows surrounding these loci. Nevertheless, a cluster of SNPs upstream of the *FADS* region in maritime hunter-gatherers exhibited values above the 1^st^ percentile threshold (*see Supplementary Note 9, Figure S9-10*), yet this signal was entirely absent in the 36 ancient samples from Patagonia and Tierra del Fuego.

Subsequently, we examined selection signals within the top 1% of PBSn1 values across both ancient and masked genomes of PHG and MHG descendants (*see Supplementary Note, Figures S11-12*). In PHG individuals, 14 SNPs overlapped with signals in ancient genomes, predominantly associated with genes involved in immunological processes (*CREG1*, *RCSD1*, *TRIM3*, *ZFPM2*) (Astle et al., 2016; Wojcik et al., 2019), skeletal and height-based phenotypes (*MNAT1*, *SLC4A4*, *TIAM2*, *LRP12*) (Yengo et al., 2018), and body mass traits (*NRXN1*, *NRG1*, *WRN*) (Astle et al., 2016; Pulit et al., 2018). For MGH, we identified 21 shared SNPs, mostly associated with height (*AGMO*, *CARD11*, *XKR6*, *MTMR9*, *DENND3*, *CDCA2*) (Kim, 2018; Yengo et al., 2018; Watanabe et al., 2019), immunological processes (*ACTR10*, *ACOXL*) (Astle et al., 2016), and metabolic processes (*GXYLT1*, *C1RL*, *PTK2*) (Kanai et al., 2018; Watanabe et al., 2019).

### Functional annotation of genes in loci of selection signatures

We further identified phenotypes associated with the strongest selection signals from PBSn1 and iHS tests using the Atlas of GWAS Summary Statistics (https://atlas.ctglab.nl/). Among the 39 genes from the top 10 genomic regions in PHG, 12 (30.8%) were linked to height-related traits, with *CADM1*, *RERE*, *SUPT3H*, *CRIM1*, and *RUNX2* exhibiting the strongest GWAS association signals (p-values 10^-44^) (Kim, 2018; Yengo et al., 2018). Additionally, nine genes (23.1%) were associated with celiac diseases (i.e., *TRIM31*, *TRIM36*, *TRIM15*, *TRIM10*, *TRIM40*, *RNF39*, *PPP1R11*, and *ZNRD1*) (Trynka et al., 2011), while seven genes (17.9%) were linked to metabolic traits, including body mass index, weight, impedance measures, trunk fat percentage, phosphatidylcholine levels, and Omega-3 fatty acid metabolism (*RGS17*, *TMEM161B*, *PTPN13*, *NSMCE2*, *ME3*, *TCEA1*, and *LYPLA1*) (Kettunen et al., 2016; Pulit et al., 2018; Watanabe et al., 2019).

For MHG descendants, key genes identified through selection scans were associated with metabolic trait variation (Supplementary Table S14). Notably, 12 out of 30 selected genes (40%) were linked to lactate dehydrogenase blood levels (*CD163L1*, *ACSM4*, *PEX5*), cholesterol levels (*PSRC1*, *CELSR2*, *MYBPHL*, *KIAA1325*), impedance measures (*SORT1*, *PSMA5*, *TIAM1*), hip circumference (*GABRA5*), and gamma-glutamyl transferase (*SOX17*) (Global Lipids Genetics Consortium, 2013; Kanai et al., 2018; Watanabe et al., 2019).

We conducted a comprehensive analysis of over-represented GWAS phenotypes and functionally relevant biological processes among the top 1% of PBSn1 values, encompassing 3803 SNPs spanning 1617 genes (including intronic, exonic, UTRs, ncRNA, and splicing variants) (Supplementary Table S8, S12). In PHG descendants, functional enrichment analysis was performed using the GENE2FUNC tool within FUMA (Functional Mapping and Annotation) GWAS platform (https://fuma.ctglab.nl/). This analysis identified a cluster of 11 genes (*CHSY1, UGT1AB*, *UGT1A10*, *UGT1A9*, *UGT1A7*, *UGT1A6*, *UGT1A5*, *UGT1A4*, *UGT1A3*, *UGT1A1, B3GAT2*) significantly associated with the Gene Ontology (GO) Biological Processes involved in flavonoids and xenobiotics glucuronidation as well as flavonoid metabolic process (Supplementary Table S15). Additionally, these genes were enriched for GO Molecular Functions related to glucuronosyl transferase activity (Supplementary Table S16). Furthermore, this gene cluster exhibited strong enrichment in GWAS for total bilirubin levels (Supplementary Table S17). The most significantly overrepresented GWAS phenotype was post-bronchodilator FEV1/FVC ratio, with 36 genes, including *SUPT3H*, demonstrating robust associations (Supplementary Table S17).

In the case of MHG descendants, we identified 630 genes within the top 1% of PBSn1 values (3652 SNPs). Functional annotation revealed significant enrichment for neurobiological pathways, alongside two immunological traits associated with T-cell interactions (Supplementary Tables S18-20). Strikingly, obesity-related traits exhibited the strongest enrichment signal (adjusted p-value 5,88e^-9^), with 79 of 596 genes previously linked to obesity also present in our dataset (Supplementary Table S21). Among these, *PRKG1* and *CPT1A* stood out as notable candidates, having been previously described as targets of positive selection in Siberian population (Cardona et al., 2014; Clemente et al., 2014).

Through an ancestry-based selection framework, we detected signatures of adaptation in immune system related loci both preceding and following European contact. Notably, a 3.05Mb region within the major histocompatibility complex (MHC) on chromosome 6, characterized by increased Native American ancestry, was significantly enriched for genes implicated in a range of diseases and immune-related traits. These include autism spectrum disorder, HIV-1 control, ulcerative colitis, white blood cell count, Takayasu arteritis, and inflammatory bowel disease, as evidenced by GWAS data (Supplementary Tables S22). Additional selection peaks on chromosomes 12, 15, 2, and 21 were associated with key metabolic and physiological processes, including bilirubin levels (*SLCO1B3, SLCO1B7, SLCO1B1, SLCO1A2*), age at menarche [*CCDC85A* associated in (The GIANT Consortium et al., 2010)], psychiatric disorders [*VRK2* associated in (Tesli et al., 2016)], and cancer susceptibility [*CXADR, BTG3* associated in (Nilchian et al., 2019)], respectively.

The results of genome-wide scans for positive selection before and after admixture events in Patagonian individuals are summarized in Figure 5. This general model was constructed based on both the observed genomic signals and the admixture history inferred from the populations analyzed in this study. Selection signals were categorized into three main phases: (1) those detected in the earliest populations inhabiting Patagonia between 12,000 and 7,000 yBP, primarily involving genes related to height and lipid metabolism; (2) signals specific to maritime hunter-gatherers from western Patagonia, likely reflecting adaptations to marine environments, with strong selection in genes related to lipid processing and LDL regulation; and (3) signals enriched in the HLA region on chromosome 6, suggesting immune-related adaptations to pathogen exposure following changes in the environmental and demographic landscape. Together, these findings provide a history of adaptive processes, from the earliest Native American individuals that migrated through Beringia, the first settlers of Patagonia to recent selection after admixture with Europeans.

**Figure 5.**
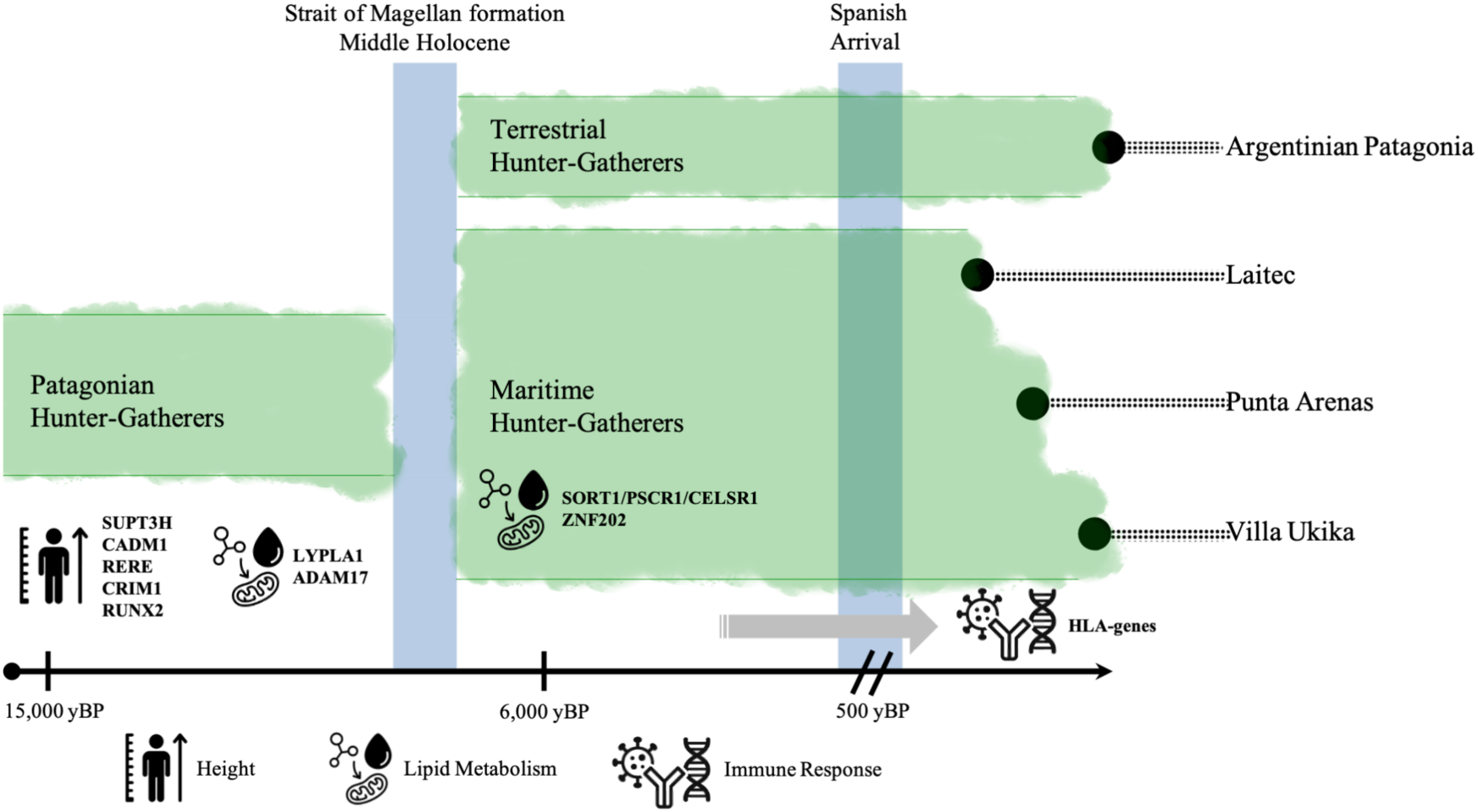
Temporal model of natural selection in Native Patagonians. Illustration of the top candidates genes under positive selection across three key phases in Patagonian population history, based on results from *PBSn1*, *iHS*, and ancestry enrichment analyses. Candidate genes are grouped into major functional categories: height regulation, lipid metabolism, and immune response. Two major demographic transitions frame these phases: the formation of the Magellan Strait and the arrival of the Spanish. The model is structured into: (1) the initial colonization and early occupation of Patagonia (prior to the formation of the Magellan Strait), (2) the emergence and diversification of maritime hunter-gatherers groups in western Patagonia, and (3) the admixture phase following European contact, marking the formation of modern admixed populations. Admixture onset (black dots) for each population are positioned according to estimates inferred using *TRACTs*. The horizontal gray arrow represents the potential contribution of pre-admixture adaptive haplotypes, retained in the Native ancestry of Maritime hunter-gatherers descendants, to post-admixture selection.

## Discussion

This study leverages novel genome-wide genotyping data from present-day Patagonians to elucidate their genetic structure, relationships with Native American groups, and signatures of local adaptations. Our findings reveal variable levels of European ancestry among all sampled individuals, except for two from Laitec, underscoring the pervasive genetic impact of European colonization across Patagonia and Tierra del Fuego. Although European incursion into the region began in the northern Patagonia in the 16^th^ century and progressively extended southward by the 20^th^ century (De Cortes Hojea & De Goicueta, 1879; Ladrillero, 1880; Hanisch, 1982), our findings suggest an earlier onset of admixture in Patagonia and Tierra del Fuego compared to other Chilean regions. Specifically, the admixture in this southern area appears to have commenced prior to that observed in central Chilean populations, where admixture events have been estimated to begin between 10 to 14 generations ago (∼1850-1650 AD) (Eyheramendy et al., 2015; Homburger et al., 2015). In Laitec, admixture is inferred to have begun around ∼1525 AD (19 generations ago), aligning with historical records of explorers, whalers, and sealers arriving by 1526, followed by the establishment of settlements such as Castro in the Chiloé Archipelago (1567) and Punta Arenas in Southern Patagonia (1848) (Emperaire, 1958; Hanisch, 1982; Martinic, 2004). In contrast, in Argentinean Patagonia, admixture appears to have initiated around (∼1774 AD, 9 generations ago), coinciding with historical narratives that describe a lack of permanent European settlements until the region was annexed by Argentina in the late 1880’s (Parolin et al., 2019). Although the gene flow from Laitec to Yámana inferred by Treemix cannot be confidently attributed to either pre- or post-European contact, the overall patterns of migration–corrborated by both Treemix and TRACTs–support historical movements from Chiloé to the Magellan region between the 16^th^ and 18^th^ centuries, culminating in effective colonization in 1843, which involved admixed populations from Chiloé (Jorquera & Jaramillo, 2020).

The Native American genomic component across Patagonia exhibits a distinct north-south division. Patagonians from Argentina and the Chilean coast, spanning from the Chiloé Archipelago (Laitec) to Kawésqar-inhabited islands near the Strait of Magellan, shared ancestry with the Mapuche population. In contrast, the Yamana of Chilean Tierra del Fuego display a unique genetic profile, aligning with ancient individuals from both the northern and southern coasts of the Strait of Magellan (aPatagonia and aFuegians, respectively), as previously reported in a smaller sample (de la Fuente et al., 2018). These findings underscore the complex interplay of migrations, isolation events, and genetic differentiation processes that has shaped the genetic landscape of Patagonian populations, mirroring broader continental patterns (Raghavan et al., 2015; Willerslev & Meltzer, 2021; Santos et al., 2022).

Our analyses using *PBS* and *iHS* reveal genomic signatures of positive selection in present-day Patagonians. Although the precise timing of selection events in Patagonia remains uncertain, previous studies using PBS and iHS suggest that selection time windows can range from ∼3,000 to ∼20,000 yBP (Fumagalli et al., 2015; Voight et al., 2006). These estimates align with major demographic transitions in the region, including the initial peopling of Patagonia and adaptation to cold environments (∼12,000 yBP) (Borrero et al., 1998) as well as the transition to a maritime subsistence strategy (∼7,000 yBP) (Rivas et al., 1999; Yesner et al., 2003). The identified selected regions show significant enrichment for genes involved in metabolic processes, height, and immunological response. Notably, we identified a strong selection signal in a region encompassing the *SUPT3H* gene on Native American chromosomes, previously linked to height in African Pygmies (Mendizabal et al., 2012). Additionally, we identified selection on *ADAM17*, a gene implicated in lipid metabolism (Junyent et al., 2010) and convergent adaptation for light skin pigmentation in Asian populations compared to Europeans (Norton et al., 2006). Most of the top selection signals involve height-associate genes (*SUPT3H*, *CADM1*, *RERE*, *CRIM1*, and *RUNX2*). Height variation is well-documented across diverse human populations (Allen et al., 2010; Yang et al., 2010), with Native Americans generally exhibiting shorter statures (Ruiz-Linares et al., 2014). Historical descriptions characterize Tierra del Fuego inhabitants as short, robust, and with short limbs-traits akin to Arctic populations adapted to cold environments (Pearson & Millones, 2005). Such morphology is hypothesized to minimize heat loss by optimizing volume-to-surface area ratios (Brown, 2012). Interestingly, the extinct Tehuelche population from the Northern Argentinian displayed tall stature, contrasting with this adaptive trend (Hernández et al., 1998; Béguelin, 2011; Millan et al., 2013). These findings underscore the adaptive significance of height in Patagonian populations, highlighting its role in modulating phenotypes to cope with the region’s extreme environmental conditions.

The Native American component of Patagonian genomes exhibits selection signatures associated with lipid homeostasis and energy metabolism. Genes such as *MSMO1*, *ADAM17/IAH*, *LYPLA1*, and *ABCB10* are linked to obesity risk, fatty liver disease, and lipid metabolism in response to high-fat diets (Liu et al., 2009; Junyent et al., 2010; Kobayashi et al., 2016; Suzuki et al., 2016; Seguin et al., 2017; Martinez et al., 2020). Additionally, genes involved in flavonoid metabolism, which influence lipoprotein oxidation and triglycerides regulation, are overrepresented (Peluso, 2006; Murota et al., 2018; Fardoun et al., 2020). These lipid-related traits are particularly pronounced in MHG descendants from Patagonia, with the *SORT1/PSCR2/CELSR2* gene cluster strongly associated with LDL cholesterol regulation (Arvind et al., 2014; Y.-J. Zhou et al., 2015). Furthermore, genes such as *ZNF202* are linked to HDL levels, coronary ischemic disease, lipid processing, diabetes, and obesity (Schmitz et al., 2004; Stene et al., 2008; Razzaghi et al., 2012). Obesity-related traits are the most overrepresented GWAS hits among the top 1% distribution of PBS values, including genes such as *PRKG1* and *CPT1A*, previously identified as cold adaptation targets in Siberian populations (Cardona et al., 2014; Clemente et al., 2014). These findings suggest the pivotal role of these genomic regions in shaping lipid-metabolism and cold adaptation in Patagonian individuals.

Notably, while multiple adaptation episodes have been documented across South American ecoregions, the candidate genes identified here (e.g., *SUPT3H*, *CADM1*, *RERE*, *CRIM1*, *RUNX2, MSMO1, and SORT1/PSCR2/CELSR2*) were not detected under selection in other populations—highlighting their novelty to Patagonian adaptation. Comparative analyses further reveal distinctive features of Patagonian genomes. Unlike other Native American groups (Amorim et al., 2017), Patagonians lack selection signals in the FADS region, which regulates polyunsaturated fatty acids metabolism and is under strong selection in both Arctic populations and admixed Hispanic/Latino populations (K. M. Reynolds et al., 2023). This absence may reflect genetic drift during early American settlement or the severe genetic bottleneck following European contact (Castro e Silva et al., 2022). Ancient DNA from Patagonia similarly shows no FADS selection peaks, suggesting that cold adaptation and maritime diet-related pressures either waned before European colonization or were masked by high missingness rates in ancient genomic data. Strikingly, however, we observed a noticeable overlap between ancient and present-day Patagonians in selected loci associated with height, metabolism, and—most prominently—immune function. This persistence implies that these adaptations were critical for surviving Patagonia’s extreme environments and may have conferred resilience against both endemic and introduced pathogens.

Our findings highlight the immune system’s role in ancestry-related selection following admixture. A notable discovery is a large genomic region on chromosome 6, enriched for Native American ancestry, encompassing the HLA region. While it has been suggested that European ancestry could provide greater resistance due to pathogen exposure (Wolfe et al., 2007), our results indicate that beneficial Native American haplotypes have persisted in present-day Patagonia populations. This suggests that pre-Columbian populations in Patagonia had already undergone selection in response to endemic pathogens, leaving a detectable footprint. This phenomenon of immune selection in admixed populations is not unique to Patagonia. Similar observations have been made reported in other Latin American populations with Native American or African ancestry (Deng et al., 2016; Lindo et al., 2016; Q. Zhou et al., 2016; Harrison et al., 2019; Lindo et al., 2018; Lopez et al., 2019; A. W. Reynolds et al., 2019; Caro-Consuegra et al., 2022). These findings collectively suggest that the immune response has been a dominant selective force during the past 500 years of admixture, shaping the genetic landscape of these populations. By contrast, we found no evidence of selection for European haplotypes on genes for immune response, which would be expected under a scenario of selection after admixture against pathogens brought by Europeans conquistadors.

Despite its novel insights, our study has limitations. The small sample size of Patagonia hunter-gatherers’ descendants constrains the generalizability of our findings. Furthermore, genetic diversity is influenced by factors such as genetic drift during admixture and potential sex-based outcomes, as admixture primarily involves native women. Further research incorporating additional ancient genomes would be critical for validating selection signals linked to early European contact. Moreover, the absence of phenotypic data prevented direct assessment of these selection signals’ effects in present-day populations. Finally, *iHS* was affected by the masking process due to haplotype fragmentation, yet it proved to reinforce the detection of the region containing the *SUPT3H* gene.

This study enhances the understanding of the Native American genetic ancestry in present-day Patagonians, contributing with the recovery of valuable information for local Indigenous populations seeking to re-construct their identity. Genomics research involving Indigenous communities must be conducted with a special sensibility to those populations’ vision and history. This is especially important for native groups who have been thought to be extinct (e.g., Selk’nam, Kawésqar), or their self-recognize present-day individuals to their cultural and genetic inheritance (Silva et al., 2022). The samples analyzed in this work were part of historical collections from recruitments carried out more than 30 years ago. Therefore, returning results to participants was not feasible. As an alternative, the corresponding authors approached a relevant community in southern Chiloe and organized an outreach event in 2015. Participants included local representatives of Native peoples and healthcare professionals, school students, and teachers. They returned the major findings from the study in the form of oral talks and infographics, as well as an installation of replicate of a Chono skeleton that was previously recovered from that site. The event was showcased in local media in 2015 (Mundaca, n.d), and the educational material and replica remain in place until the present, cared for by the community members, who deem them part of their patrimony. Also, Argentinian communities from the province of Chubut were actively involved in this research through a scientific-cultural transfer workshop knowledge focused on genetic science (resolution number 281/2022 S DDHH GCSG).

## Conclusion

This work presents a thorough study of the genetic composition and adaptive strategies of contemporary populations in Patagonia, elucidating the intricate historical and evolutionary dynamics that have influenced them. Our findings suggest an earlier-than-expected onset admixture in Tierra del Fuego, likely driven by early European explorers, followed by more recent migration waves, possibly of admixed individuals from Chiloé (Martinic, 1999). In conclusion, we identify a pronounced genomic differentiation along a north-south axis among Native American groups in Patagonia, with evidence of historical migrations shaping the genetic landscape.

Importantly, our results disclose notable genetic adaptations associated with lipid metabolism and height, crucial for surviving in the region’s cold climate and subsisting on a maritime diet. These adaptations, such as the robust selection signals in genes related to lipid homeostasis (*MSMO1*, *LYPLA1*, and *SORT1/PSCR2*) and height (*SUPT3H*, *CADM1*, and *RUNX2*), reflect the critical role of metabolism and body morphology in minimizing heat loss and optimizing energy use in harsh environments. Additionally, our analysis revealed compelling evidence for immune selection. We detected selection signals both before and after European contact, with pre-contact selection possibly shaped since the initial occupation and settlement of Patagonia. Notably, many of the strongest immune-related selection peaks, particularly within the HLA region, appear to have been initially adapted to local pathogens but remained under strong selection pressures following European arrival. This finding underscores the profound impact of infectious diseases on Patagonia populations over time.

Furthermore, all identified selection regions represent novel candidate loci for positive selection, as they have not been previously reported in other Native populations across South America. This suggests that these adaptive events may be unique to Patagonia, potentially driven by the region’s extreme environmental pressures. However, the possibility of convergent evolution cannot be overlooked, as we observed selection on the *SUPT3H* and *ADAM17* loci, where others have independently identified adaptative signatures in very different geographies for body high and skin color, respectively. Our research demonstrates that current admixed populations in Patagonia retain genetic continuity with original settlers who accumulated local adaptations to this remote region, which in turn were beneficial even after admixture with European conquistadors. Therefore, our study both provides insights into the continuing influence of Native American ancestry in the biology of modern Patagonian populations and provides information about characteristics of their ancestors that could not be learned from written history.

## Materials and Methods

### Samples and Genotyping

Fifty samples from Laitec and the Argentinian Patagonia were sampled (Supplementary Table S1). Chilean Patagonians from Laitec were recruited during the 1990’s as part of previous projects at the Human Genetics Program of the University of Chile and provided informed consent for the utilization of their genetic material in research. The use of these samples was approved by the Ethics Committee of the Faculty of Medicine, University of Chile, from Project CONICYT USA2013-0015: “Genomic investigation of the human biodiversity in the pre-Columbian and contemporary Chilean Patagonia.” (Resolution 116-2012). Individuals from Punta Arenas (Kawésqar) and Villa Ukika (Yámana) underwent genotyping in a prior study (de la Fuente et al., 2018). Participants from Argentina were enlisted and genotyped using panels of 99 and 46 Ancestry Informative Markers (AIMs) (Avena et al., 2012; Parolin et al., 2019). Argentinean Patagonians were recruited under the supervision of María Laura Parolin and approved by the Ethics and Teaching Committees of the Sub-zonal Hospital of Esquel, Regional Hospital of Comodoro Rivadavia, Zonal Hospital of Trelew and Andrés Isola Zonal Hospital of Puerto Madryn (Resolution 009/2015). These participants consented for genomic studies and were informed about their birthplace and that of the two preceding generations (parents and grandparents) through a questionnaire. The criteria for inclusion of individuals in the present manuscript encompassed having all parents and grandparents born in Chubut Province, having the largest Native American ancestry in the sample as estimated by the AIMs and by having a haplogroup of Native American origin in chromosome Y and mitochondrial DNA. Based on these criteria, three individuals were excluded due to having direct relatives from Chile and Bolivia.

DNA samples were genotyped with the Axiom LAT1 microarray (World Array 4, Affymetrix Inc, Santa Clara, CA) in the Institute for Human Genetics, University of California, San Francisco, California, and called against the GRCh37/hg19 assembly, for a total of 585,215 SNPs distributed homogeneously throughout the genome. A total of 868 SNPs were removed as quality control parameters (missing genotype rate (>5%), identity by descent, and Hardy-Weinberg equilibrium (10^-6^)) from *PLINK v1.9* (Chang et al., 2015) (see Supplementary Note 4, Supplementary Table S2).

### Genetic Structure and Admixture Dynamics

The dataset was merged with individuals from Africa (n=30), Europe (n=30), and Asia (n=30) that were genotyped by the 1000 Genomes phase3 (Auton & Salcedo, 2015). Furthermore, for this and subsequent analyses, we incorporated a South American panel comprising 20 populations (Lindo et al., 2018; Reich et al., 2012) (https://reich.hms.harvard.edu/datasets), alongside 36 ancient representatives from Patagonia and Tierra del Fuego, spanning the middle and late Holocene. These samples occupied the geographic areas from which the contemporary samples were collected. The ancient Kawésqar, and Aonikenk populations were categorized as ancient Patagonia (aPatagonia), while the Yámana, Haush, Selk’nam populations were categorized as ancient Fueguians (aFueguian) (Raghavan et al., 2015; de la Fuente et al., 2018; Moreno-Mayar et al., 2018; Nakatsuka et al., 2020; Mallick et al., 2023). The comprehensive list of reference populations is presented in Supplementary Table S3 and Supplementary Note 6. A total of 35,788 SNPs were identified as common with the reference dataset. A principal component analysis (PCA) was conducted using *smartPCA*, as integrated within the *EIGENSOFT* program (Patterson et al., 2006). To characterize the ancestral components within the sample, we employed the *ADMIXTURE* (Alexander et al., 2009) software. SNPs with minor allele frequencies (*MAF*) less than 1% and under linkage disequilibrium (*LD*) were excluded using *PLINK v1.9* (Chang et al., 2015). The software was run assuming 2 to 10 “ancestral” populations (K = 2 to K = 10) in 10 replicates and plotted using *Pong* software (Behr et al., 2016). We used a cross-validation test to evaluate the fit of each K. Relatedness among samples was estimated by *REAP* (Thornton et al., 2012) through pairwise identity-by-descent (IBD) comparison, where one individual from each pair with a threshold > 0.125 was excluded. *TreeMix v1.13* (Pickrell & Pritchard, 2012) was performed to detect population divergences and directional migration events, employing maximum likelihood estimations in Patagonia and the reference populations. A range of 0-10 migration events was evaluated with their respective residual and likelihood values. We phased the dataset using *SHAPEIT2* (Delaneau et al., 2012), referencing the 2,504 individuals from the 1000 Genomes project phase 3. Subsequently, we inferred the local ancestry of chromosomal segments via *RFMix v1.5.4* (Maples et al., 2013) and categorized each segment according to its genetic origin as European (IBS, n=30), African (YRI, n=30) or Native American (MAP, n=34). *TRACTs* (Gravel, 2012) analysis was conducted to determine the onset of the admixture process and potential migration pulses in accordance with different demographic models. We assessed the likelihood of four distinct scenarios: i) a single pulse involving Europeans and Native Americans (EUR+NAT); ii) a continuous pulse associated with either Europeans (EUR+NAT, EUR) or Native Americans (NAT+EUR, NAT); iii) two pulses related to the initial encounter and subsequent migration for additional ancestry (e.g. EUR+NAT, AFR); and iv) three pulses assuming two different migrations following the initial interaction (e.g., EUR+NAT, AFR, EUR). The feasible combinations for ancestry participation are available in *Supplementary Figure S7*. Likelihood values were compared for each migration model utilizing Bayesian information criteria (BIC). We remove related individuals (*REAP*) or those with ancestry proportions above 95% of a singular continental population (European, Native American, or African) as these individuals either represent non-admixed individuals or exceedingly recent migrants. Following these filters, we retained a total of 18 individuals from Laitec, 14 from the Argentinian Patagonian, 3 from Kawésqar, and 13 from Yámana.

### Admixture-based selection

To investigate the signatures of selection linked to ancestral components, we scanned a comprehensive examination for loci exhibiting a significant enrichment of Native, European, or African ancestry (Bhatia et al., 2014). For this analysis, we evaluated the mean global ancestry in relation to the local ancestry at each Single Nucleotide Polymorphism (SNP), including all individuals from Patagonia, and the resulting comparisons were visualized using R (R Core Team & Team, 2014). The genome-wide significant signal of selection as a deviation in local ancestry was analyzed through a Z-test comparison at each locus. A one-tailed p-value of less than 5% was established as statistically significant.

### Pre-European based selection

In order to mitigate the potential for ancestry misinterpretation concerning selection, we executed a Github R-script repository designed to obscure all non-native ancestry segments (African and European) within the genomic composition of each individual (https://github.com/PatricioPezo/Masking_Ancestry). Due to the substantial volume of missing data resultant from the masking procedure, we used the option *–vcf-half-call m* to facilitate the conversion from VCF to Plink formats (see Supplementary Note 5). As a first approach, we used a modified method to the Population Branch Statistic (*PBS*) (Yi et al., 2010), which calibrates the statistic and avoids the occurrence of artificially elevated PBS values in the context of lower or higher differentiation among all populations (Crawford et al., 2017). *PBSn1* was conducted for two distinct groupings of individuals from Patagonia: a) Patagonian Hunter-gatherers (PHG), encompassing the ancestors of individuals from Laitec, Argentinian Patagonia, Kawésqar and Yámana, and b) Patagonian Maritime Hunter-gatherers (MHG), which includes the ancestor of individuals from Laitec, Kawésqar, and Yámana. *PBSn1* operates on the basis of pairwise comparisons utilizing the Fst coefficient among three populations: the target group, an outgroup and a closely related genetic group not affected by the same selective pressure. In this context, for the PHG, we designated East Asians as the outgroup, while the Mapuche from south-central Chile were considered as the closely related genetic group (EAS: MAP: PHG). In the case of MHG, we incorporated Argentinian Patagonia as the closely related genetic group and East Asian as the outgroup (EAS: ARP: MHG) (*see Supplementary Note 11, Figure S13*). The integrated haplotype score (*iHS*) was likewise implemented using *selscan v3.2* (Szpiech & Hernandez, 2014) on the masked data to facilitate a comparison with the findings obtained from *PBSn1*. *iHS* values exceeding 2.5 were classified as candidates for selection, corresponding to the upper 1% tail of the distribution (Coop et al., 2009). In light of the considerable quantity of missing data due to the European-masking process within our sample, we retained 16 individuals with at least 77% of native ancestry to ensure the preservation of the maximum haplotypes for *iHS* analysis purposes. Both *PBSn1* and *iHS* were computed for each SNP utilizing 200 Kb non-overlapping windows (Coop et al., 2009; Voight et al., 2006) through *bedtools* (Quinlan & Hall, 2010), while adhering to a minimum threshold of 20 SNPs.

### Variant Annotation, Biological processes, and GWAS comparison

Potential genetic candidates for selection associated with *PBSn1*, *iHS*, and Ancestry enrichment were systematically analyzed. Gene annotation was conducted using *Annovar* (Wang et al., 2010) through the “Known Canonical Genes” database in USCS Genome Browser (Hsu et al., 2006). Subsequently, we conducted a search for phenotypic associations reported in Genome-wide association studies (GWAS) to evaluate the biological significance of our selected variants via the GWAS Atlas database (Watanabe et al., 2019) (https://atlas.ctglab.nl). The enrichment analysis of GWAS and biological processes was performed on the genomic regions under selection in the sample utilizing the Functional Mapping and Annotation of Genome-Wide Association Studies (FUMA GWAS), employing the *GENE2FUNC* function, with 20,434 utilizing as the reference gen set, *Ensembl v.102*, and applying the Benjamini-Hochberg correction for gene-set enrichment testing (FDR < 0.05) (Watanabe et al., 2019).

## Data Availability

Individual-level genotypes for new data presented here are available upon request through a data-access agreement with the University of Chile. Access requests should be directed to the corresponding authors by using the form provided in Supplementary Material online, Text S1.

## Acknowledgments

We would like to thank all Patagonian individuals and the local communities sampled who participated in the study. Also, we are grateful to Alejandro Sepúlveda and Alejandro Blanco for laboratory assistance and bioinformatic support. This work was supported in Chile by the National Agency of Development and Investigation (ANID) grants CONICYT FONDECYT 1140544, USA2013-0015, and FONDEQUIP EQM140157 and by the National Agency of Technologic and Scientific Promotion (PICT) 2013-2014 in Argentina.

