## Supplementary Information for "Present-day admixed genomes reveal prehistoric adaptation to cold, maritime diet, and local pathogens in Patagonia"

**Author Contributions: Patricio Pezo**: Investigation, Writing - Original Draft; Formal Analysis; Visualization. **Michael Orellana-Soto**: Formal Analysis; Visualization. **Alexandra Salcedo**: Formal Analysis; Visualization. **Juan Esteban Rodríguez-Rodríguez**: Formal Analysis. . **Cristian Yáñez**: Formal Analysis. **Elena Llop:** Investigation. **Eugenio Aspillaga:** Investigation. **María Laura Parolín**: Supervision; Writing – Review & Editing. **Andrés Moreno-Estrada**: Conceptualization; Supervision; Resources. **Carlos Bustamante**: Conceptualization; Resources. **Julian R. Homburger**: Formal Analysis. **Christopher R. Gignoux**: Formal Analysis. **Alexander Ioannidis**: Supervision. **Celeste Eng:** Investigation. **Scott Huntsman**: Investigation. **Esteban G. Burchard**: Supervision; Resources. **Tábita Hünemeier**: Supervision; Writing-Review & Editing. **Mauricio Moraga**: Investigation; Supervision; Writing – Review & Editing; Resources. **Ricardo Verdugo**: Conceptualization; Supervision; Writing – Review & Editing; Project Administration; Resources.

**Competing Interest Statement**: CRG owns stock in 23andMe, Inc and JRH is currently an employee of Novo Holdings. Other authors declare no competing interest.

**This PDF file includes:**

Supplementary text
Figures S1 to S12
SI References

### Supplementary Note 1: Geographic and Environmental Overview of the Patagonia region

Patagonia is a vast and ecologically diverse region in southern South America, extending from approximately 40°S to 55°S, and encompassing Tierra del Fuego as well as the southern reaches of the Andean Cordillera. The western Patagonia margin is defined by a narrow strip of land stretching from the rugged Pacific coastline to the high Andean peaks, which exceed elevations of 1,500 meters above the sea level (Garreaud et al., 2013). In contrast, eastern Patagonia is characterized by a semi-arid environment dominated by drought-resistant grasses and shrubs, forming part of the Patagonian steppe biome (Scheinsohn, 2003). This marked environmental gradient across Patagonia has played a critical role in shaping both the ecological and cultural trajectories of human populations throughout the Holocene.

### Supplementary Note 2: Marine Hunter-Gatherer groups of Western Patagonia

Native populations of Patagonia historically followed two distinct subsistence strategies: terrestrial hunter-gatherers, who occupied the steppe regions east of the Andes, and marine hunter-gatherers or canoe-faring groups, who inhabited the forested and humid archipelagos along the western Patagonian coast (Massone et al., 2016).

These nomadic canoe-faring groups–active for at least 6,000 years–relied on marine hunting, fishing, and shellfish gathering. Over time, they developed into the historically documented Kaweskar, who inhabited the archipelagos between the Golfo de Penas and the Brecknock Peninsula, and the Yamana, located from the Beagle Channel to Cape Horn (Massone et al., 2016). Archaeological evidence suggests that throughout this period, their settlement patterns and toolkits remained relatively simple, consistent with European accounts describing their shelters, clothing, and social organization as minimally elaborated (Orquera & Piana, 2009).

Several environmental factors supported the emergence of a coastal subsistence strategy in this region: rugged terrain that limited overland mobility, abundant high-calorie marine resources along the coastline, and a scarcity of terrestrial plant and animal resources in the temperate forests (Orquera & Piana, 2009). The nearshore one provided pinnipeds, seabirds, fish, mustelids, and occasionally stranded cetaceans, as well as driftwood and lithic materials suitable for tool production (Orquera et al., 2011).

### Supplementary Note 3: Admixture Dynamics and Native American Population Decline

Following European contact, Native American populations in South America experienced a dramatic demographic collapse, with effective population sizes declining by more than 90% (Castro e Silva et al., 2022). Pre-Columbian estimates suggest the continent was home to over 24 million individuals, a number drastically reduced by warfare, forced displacement, and epidemics introduced by Europeans—including influenza, measles, and novel strains of tuberculosis (Adhikari et al., 2017; Patterson et al., 2006; Thornton, 1997; Lindo et al., 2018). For example, the Kawésqar population in southernmost Patagonia plummeted from approximately 3,000–4,000 individuals before European arrival to 500, and further declined to just 60 by 1953 (Gleisner & Montt, 2014; Martinic, 2004).

From the 17th to 19th centuries, extensive admixture reshaped the genetic landscape of South America, involving Indigenous peoples, European colonizers, and African forced migrants (Gravel et al., 2013; Homburger et al., 2015; Adhikari et al., 2017). These admixture patterns varied considerably by region, reflecting diverse colonial histories and post-contact dynamics (Gravel et al., 2013; Homburger et al., 2015). In Chile, genomic data show relatively balanced contributions from European and Native American ancestries, though Native ancestry is more prominent in Chilean Patagonia (~46%) compared to Argentine Patagonia (~35.8%) (Eyheramendy et al., 2015; Verdugo et al., 2020).

### Supplementary Note 4: Quality Control and Variant Filtering

Genotyping quality control for the Axiom Latino Array (World Array 4, Affymetrix Inc., Santa Clara, CA) was conducted using *PLINK v1.9* (Chang et al., 2015). The pipeline included filtering based on individual and SNP-level missingness, genotype error rates, Hardy-Weinberg equilibrium (HWE), minor allele frequency (MAF), sex concordance, and identity-by-descent (IBD). Specifically, SNPs with >5% missing data, individuals with >5% missing genotypes, and markers deviating significantly from HWE ($p < 1 \times 10^{-6}$) were excluded to minimize technical artifacts.

Among the 607,853 initial SNPs, 868 were filtered out based on missingness, HWE deviation, and relatedness criteria, resulting in a high-quality dataset of 584,347 SNPs distributed homogeneously across the genome (Supplementary Table S2). Only one individual—belonging to the Tehuelche group—exhibited >5% missingness and was removed. A total of 87 SNPs showed genotyping error rates above 5% and were also excluded. 782 variants significantly out of HWE were removed as potential genotyping artifacts. IBD analysis revealed 64 pairwise comparisons exceeding 12.5% genetic relatedness.

### Supplementary Note 5: Masking Non-Native Genomic Regions

To retain only Native American haplotypes in downstream analyses, we masked all SNPs inferred to carry non-Native ancestry—defined here as European, African, or Unknown—based on local ancestry inference performed with RFMix. This masking process ensured that only genomic segments confidently labeled as Native American were preserved. As shown in Supplementary Figure 6, the procedure began with phased genotype data in VCF format, generated using *SHAPEIT2* (Delaneau et al., 2012) and converted with *VCFtools* (Danecek et al., 2011). Individual-level VCF files were created using the --subset function in VCFtools.

Subsequently, each individual's maternal and paternal chromosomes were separated and annotated in R (R Core Team & Team, 2014) via an overlap join, which assigned local ancestry labels to each SNP based on the start and end positions provided in the *RFMix* output (Maples et al., 2013). All SNPs not assigned a Native American ancestry label were replaced with missing genotype values ("."). Finally, both haplotypes were recombined per individual, and a masked, phased genotype dataset was compiled for all samples. The full masking pipeline is publicly available at https://github.com/PatricioPezo/Masking_Ancestry.

**Supplementary Note 6: Native American Reference Samples**

To characterize Native American ancestry, we employed a comprehensive reference panel comprising both present-day and ancient individuals. Contemporary Native South American samples were primarily obtained from the Reich Lab database (Reich et al., 2012) and include the following populations: Arhuaco, Wayuu, Kogi, Embera, Waunana, Guahibo, Inga, Piapoco, Palikur, Ticuna, Quechua, Aymara, Chane, Wichi, Guarani, Toba, and Diaguita. Two additional populations from South-Central Chile—Pehuenche and Huilliche—were incorporated from independent genomic studies due to their geographic and cultural relevance(Lindo et al., 2018).

Ancient reference individuals were drawn from the Allen Ancient DNA Resource (AADR) (Mallick et al., 2023) and represent a temporal span from the Middle to Late Holocene. These include early samples such as Ayayema, Punta Santa Ana, and La Arcillosa2, as well as later-period individuals associated with Indigenous groups from southern Patagonia: Kawéskar and Yámana (southwestern archipelagos), and Selk'nam, Aonikenk, and Haush (southeastern mainland and Tierra del Fuego). A detailed overview of sample provenance and cultural affiliation is presented in Supplementary Figure S1 and Supplementary Table S3.

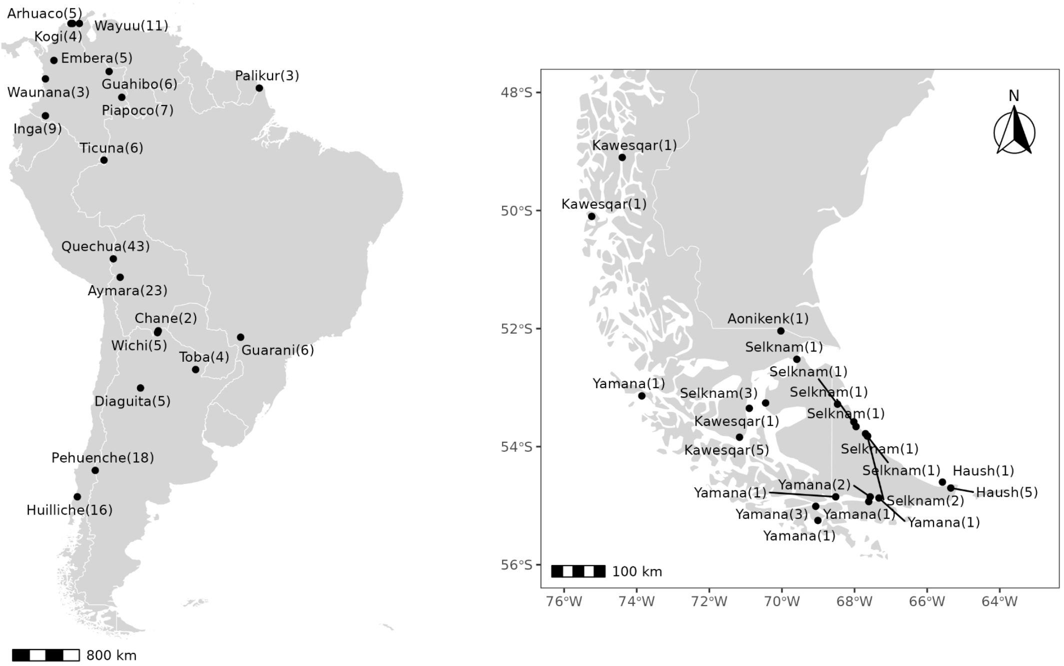


**Figure S1 | Geographic distribution of South American populations.**

Native American reference panel (left), and ancient DNA samples from Patagonia (right). South American populations and ancient samples were extracted from Reich database (Reich et al., 2012) (https://reich.hms.harvard.edu/datasets), and the Allen Ancient DNA Resource (AADR, https://reich.hms.harvard.edu/allen-ancient-dna-resource-aadr-downloadable-genotypes-present-day-and-ancient-dna-data) (Mallick et al., 2023) respectively.

### Supplementary Note 7 | Population Structure and Admixture Patterns in Present-Day Patagonians

To explore the genetic structure of four present-day Patagonian populations, we first conducted a principal component analysis (PCA) using *smartPCA* (Patterson et al., 2006) to assess patterns of genetic variation. Principal Component 1 (PC1), accounting for 5.67% of the total variance, captured continental-level differentiation by separating African and European ancestries. Principal Component 2 (PC2), explaining 2.42% of the variance, distinguished the Native American component. In the PC1–PC2 space, individuals from Patagonia are positioned between the European and Native American reference panels, highlighting their admixed genomic profile. Notably, the Kawéskar and Yámana samples showed a closer affinity to the European cluster (Supplementary Figure S2).

Next, global ancestry proportions were inferred using *ADMIXTURE* (Alexander et al., 2009), with cross-validation identifying K=5 and K=6 as the optimal models (Supplementary Figure S3). At K=4, major continental ancestries (African, European, Asian, and Native American) were clearly distinguished. Higher K values (K=6–K=9) revealed sub-continental Native American structure, separating Amazonian (e.g., Suruí, Karitiana), Andean (e.g., Quechua, Aymara), South-Central Chilean (e.g., Pehuenche, Huilliche), and southern Patagonian ancestries (e.g., ancient Kawéskar and Yámana) (Supplementary Figure S4).

Focusing on the K=6 solution and excluding highly drifted populations (e.g., Suruí and Karitiana), we observed that present-day Patagonians exhibit a dual ancestry composed primarily of Native American (74.23%) and European (24.33%) components. Within the Native fraction, ancestry is further subdivided into a distinct Patagonian component (11.55%) and a South-Central Chilean component (57.03%), underscoring the complex demographic history and regional admixture dynamics in Patagonia (Figure 2; Supplementary Table S4).

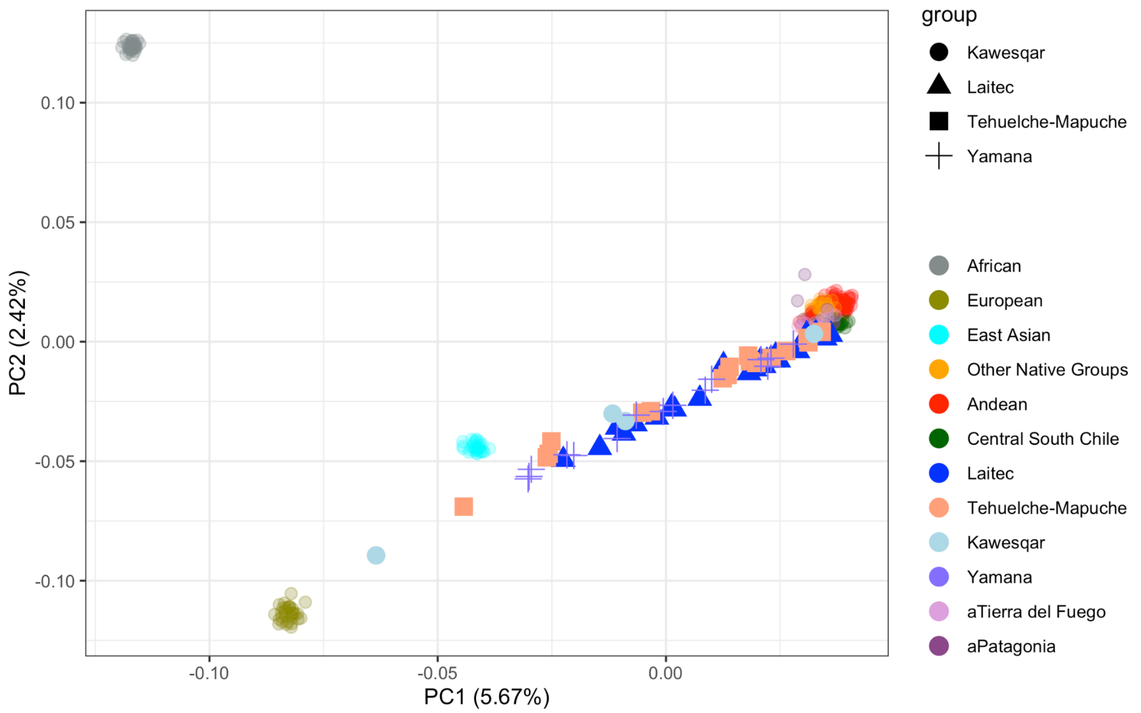


**Figure S2 | Principal Component Analysis (PCA).**

20 populations from South America (Reich et al., 2012), 36 ancient samples from Patagonia (de la Fuente et al., 2018; Moreno-Mayar et al., 2018; Nakatsuka et al., 2020; Raghavan, 2015), and a panel of African (YRI, 30), European (IBS, 30), and East Asian (CHB, 30) from 1000G (https://www.internationalgenome.org/) were analyzed. Each dot represents an individual from the following groups: Andean (Quechua, Puno, Aymara, and Diaguita), Central-South Chile (Pehuenche and Huilliche), other native groups from lowland South America (Arhuaco, Kogi, Wayuu, Embera, Guahibo, Waunana, Palikur, Piapoco, Inga, Ticuna, Chane, Wichi, Guarani, and Toba), and ancient DNA samples from Patagonia and Fueguians. The different point shapes were associated with Patagonia populations (Laitec, Kawésqar, Yámana, and Tehuelche-Mapuche).

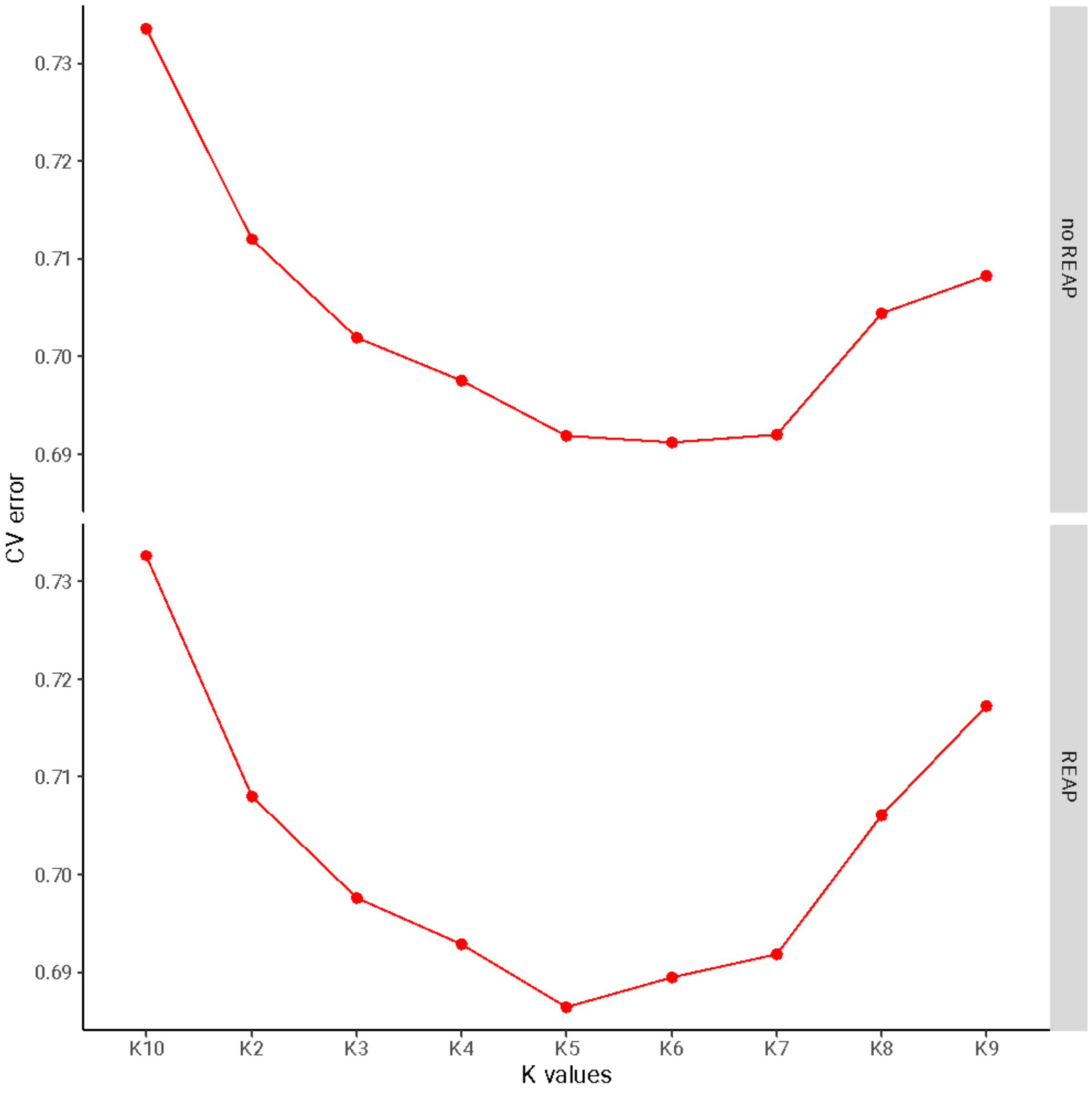


**Figure S3 | Cross validation error for each K value according to ADMIXTURE.** Likelihood distribution is grouped without (top) and with (bottom) REAP filter. The x-axis are K components (from 2 to 10), and the y-axis are Cross Validations errors.

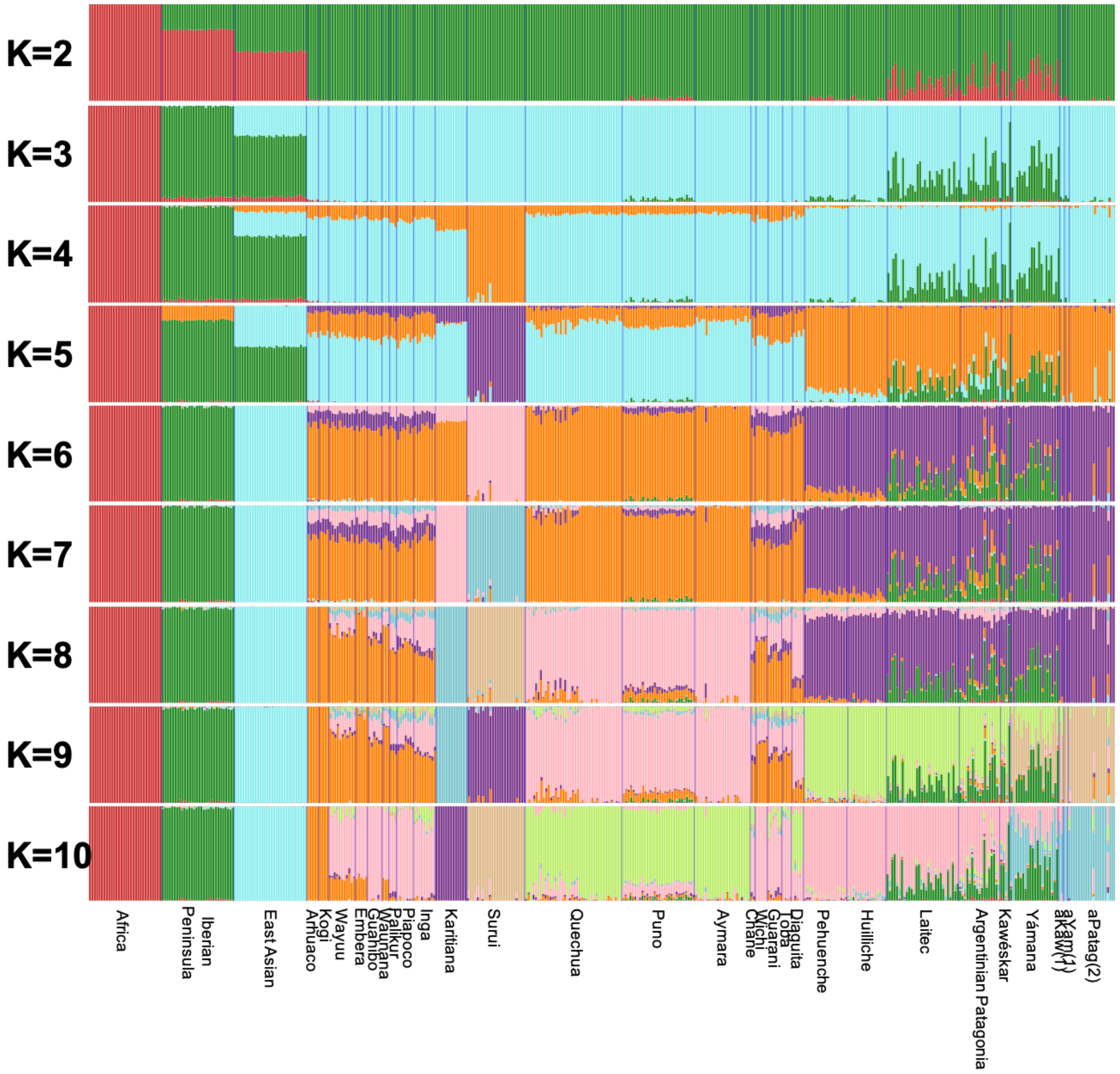


**Figure S4 | Global ancestry between Native American and Patagonia.**

Global ancestries performed by *ADMIXTURE*. Ancient and present-day northeastern Patagonians (Laitec, Argentinian Patagonia, Kaweskar, Yamana) are shown alongside reference populations including a reference panel from Africa (Yoruba), Europe (Iberian), East Asia (Beijing), and a Native American dataset (combined panel from Reich et al., 2012, and Lindo et al., 2018). The values of K's ancestral components range from 2 to 10. Each bar corresponds to each individual and the color bars represent an ancestry. Native American panels were displayed by populations (bottom) sorted by latitude (north to south).

We applied *TreeMix v1.1* (Pickrell & Pritchard, 2012) to investigate population structure and infer gene flow using allele frequency data under a maximum-likelihood framework. Ten independent models were tested by sequentially increasing the number of migration edges (m = 0 to 10), and log-likelihoods were computed for each resulting topology. Trees were rooted with the Han population to establish a directional framework. As illustrated in Supplementary Figures S4–S5, the most substantial gains in model fit occurred between m = 0 and m = 2, after which improvements plateaued, indicating an elbow-like pattern. The initial topology (m = 0) revealed clear Native American sub-structure within the Southern Cone, distinguishing groups such as Pehuenche, Huilliche, and Patagonian populations (including ancient individuals) from other South American lineages (Supplementary Figure S6).

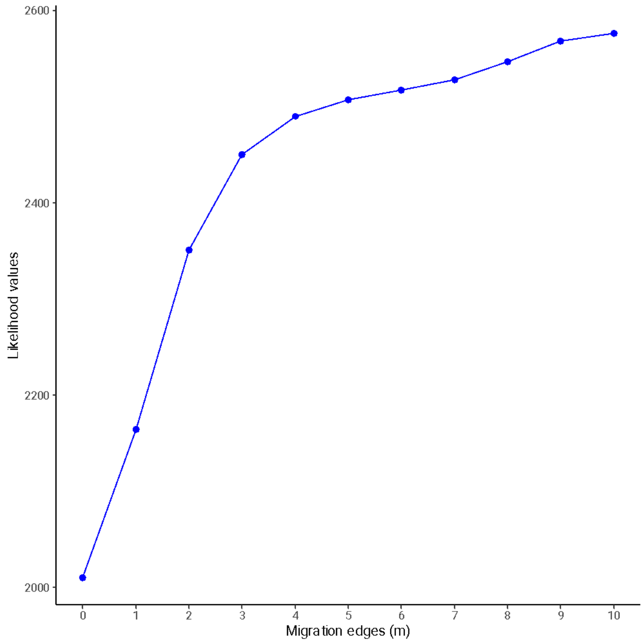


**Figure S5 | Likelihood values for each migration edge.**

Likelihood values (y-axis) across ten independent runs using Treemix v.1.13 (Pickrell & Pritchard, 2012). Each run added up one migration edge per each run (x-axis).

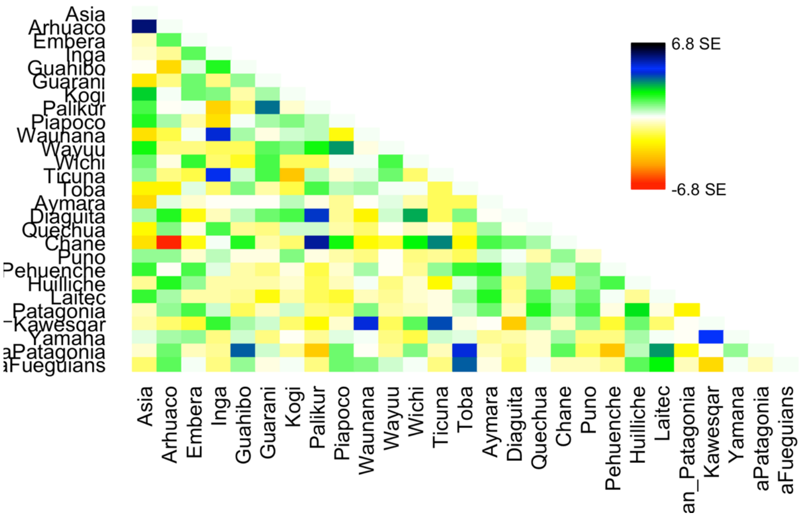


**Figure S6 | Residual fit from the maximum likelihood tree in Figure 1B with two migration edges.**

Residual covariance from each pair of populations was divided by the average standard error across all pairs. Colors are described in the palette on the right. Residuals above zero represent populations that are more closely related to each other than in the best-fit tree and thus are candidates for admixture events.

### Supplementary Note 8: Admixture Timing and Migration Dynamics in Present-Day Patagonian Populations

We inferred the timing of admixture onset and subsequent migration pulses in present-day Patagonian populations using the program *TRACTs* (Gravel, 2012). Genotype phasing was performed with *SHAPEIT2* (Delaneau et al., 2012), followed by local ancestry inference using *RFMix v1* (Maples et al., 2013), incorporating Yoruba, Iberian, and Native American reference panels. *TRACTs* models the distribution of ancestry-specific tract lengths across the genome to infer the number and timing of admixture events by simulating demographic scenarios and evaluating model fit using log-likelihood and Bayesian Information Criterion (BIC).

We tested multiple models of increasing complexity (Supplementary Figure S8). The simplest model, a single-pulse admixture (*pp_model*), assumes one historical admixture event between Native American and European ancestries (Supplementary Figure S8A). A more complex model (*ppx_xxp*) simulates an initial admixture between two ancestries, followed by a later pulse from a third ancestry, with permutations of the ancestry order to test different hypotheses (Supplementary Figure S8B). Finally, an even more complex model (*ppx_xxp_pxx*) includes two successive post-admixture migration pulses, again tested with alternate configurations of ancestral sources (Supplementary Figure S8C). Each model was evaluated based on its likelihood and penalized for overfitting using BIC (Supplementary Table S6).

The best-fitting models for each population are shown in Supplementary Figure S9. The single-pulse model best explained the admixture profiles of individuals from Argentinian Patagonia and Kawésqar, as evidenced by the lowest BIC scores and congruence between observed and expected tract length distributions. However, in the Kawésqar, the fit may be limited by a small sample size (n = 4). In contrast, individuals from Laitec and Yámana populations were better modeled by the dual-migration model, involving two post-admixture pulses—one from African and another from Native American ancestry.

The presence of African ancestry in Laitec and Yámana is notable, especially given the limited historical documentation of African migration into southern Chile (Eyheramendy et al., 2015; Verdugo et al., 2020). This component likely reflects either admixture with already admixed individuals from other regions or residual African ancestry introduced during colonial times via Spanish soldiers. Similarly, the secondary Native American pulse could reflect continued gene flow from neighboring Indigenous groups post-European contact or the movement of admixed individuals carrying Native ancestry.

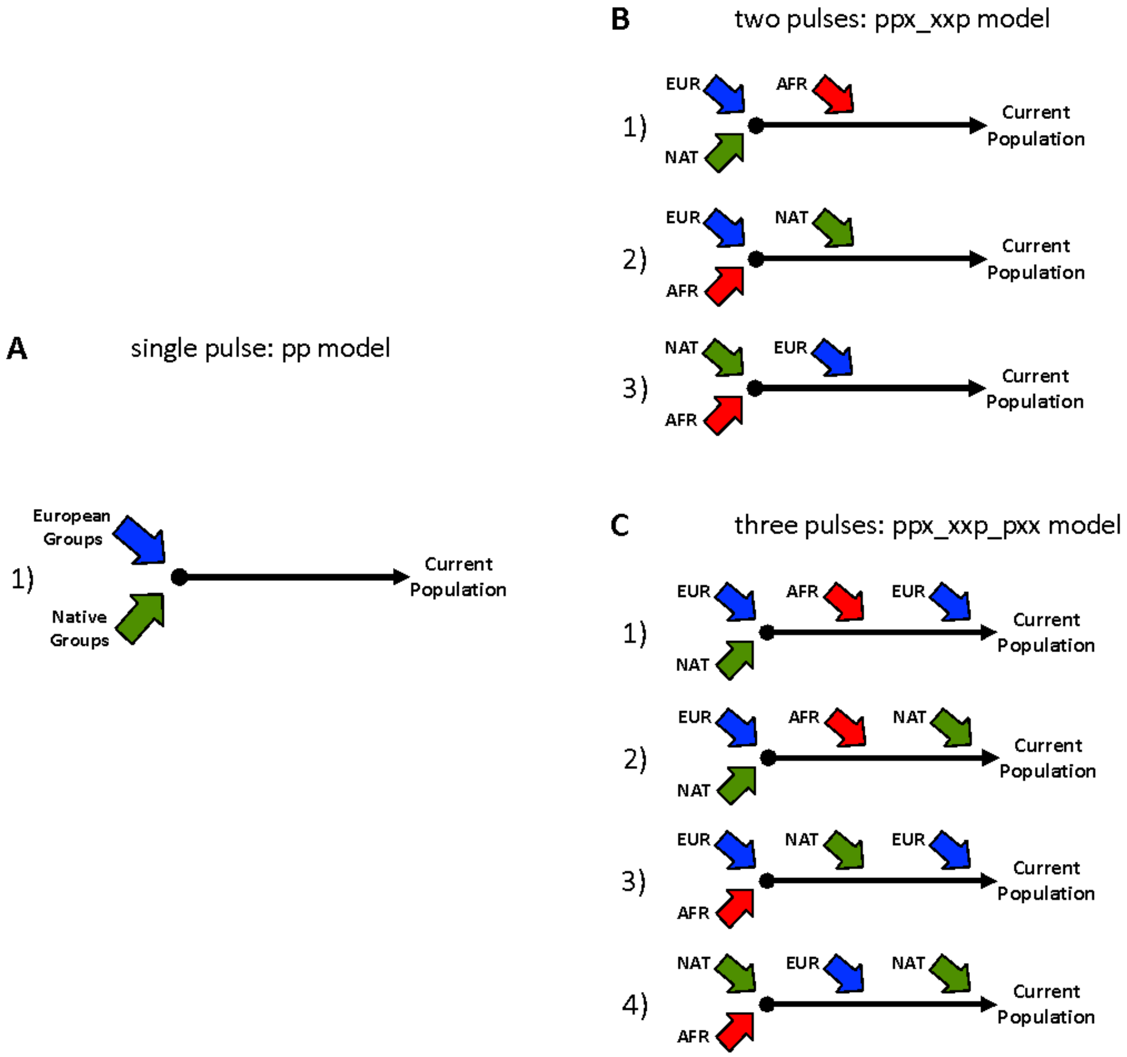


**Figure S7 | Simulation models used for TRACTs analysis.**

(A) model with a single pulse between European (blue arrow) and Native American (green arrow). B) model with a continuous pulse involving an admixture between European and Native American. We alternate another European (B1) or Native American (B2) pulse into the model. C) two pulses model associated to three different scenarios: 1) initial admixture between European (EUR) and Native American (NAT) with a subsequent African (AFR, red arrow) pulse, 2) initial admixture between European and African following a Native American pulse, and 3) Native American and African admixture following an European pulse. D) three pulses model in four different scenarios: 1) European and Native American admixture associated with two posterior African and European pulses, 2) Native American and European admixture following an African and Native American pulses, 3) European and African admixture with two subsequent Native American and European pulses, and finally 4) an initial admixture between Native American and African groups with two posterior European and Native American pulses.

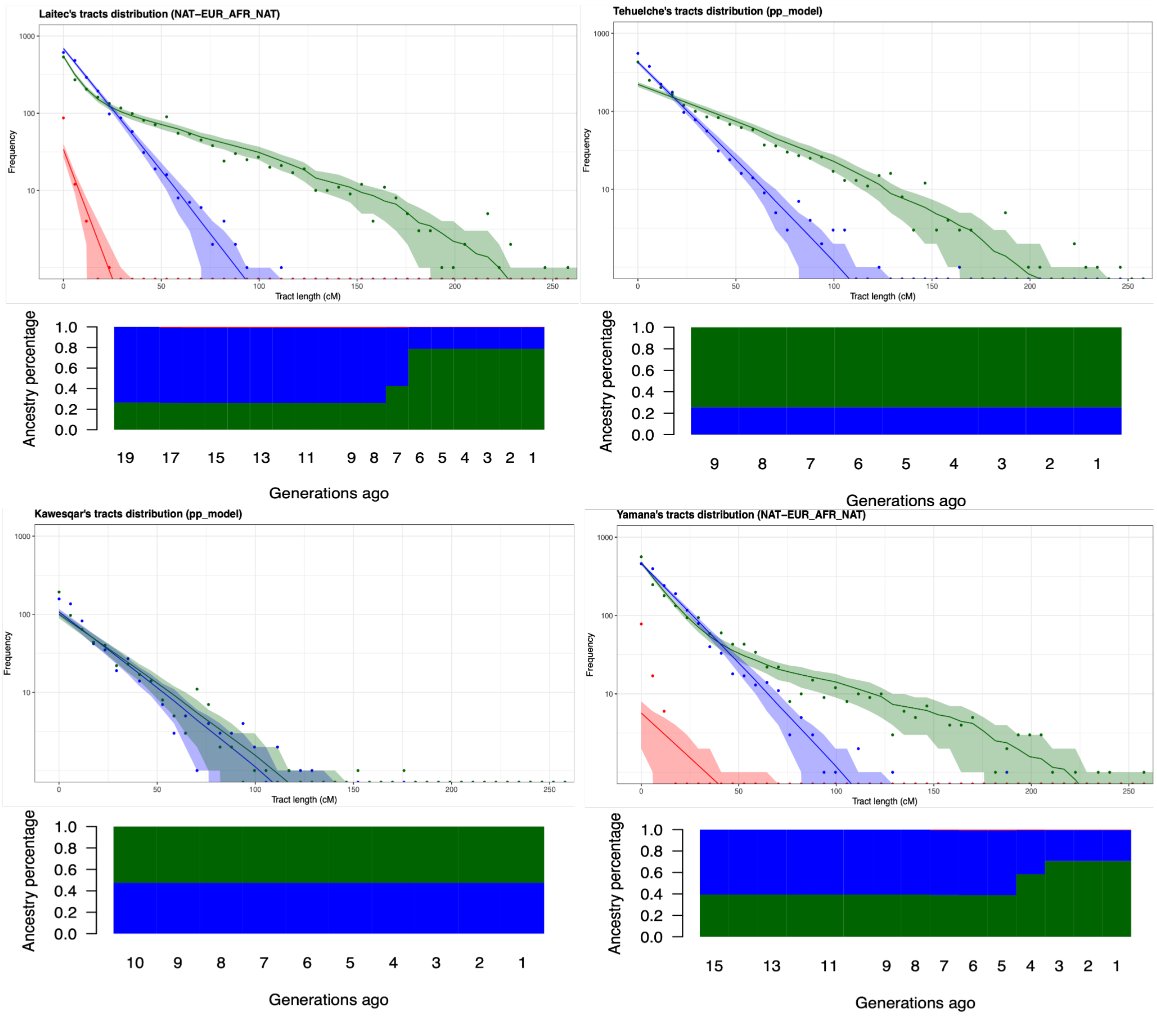


**Figure S8 | Tract length analysis and admixture times for Patagonia populations.** Both Laitec and Yamana have an initial admixture event between Native American (green) and European (blue) populations with a subsequent African and Native American pulse according to the best-fitting model. Meanwhile, Kawésqar and Argentinian Patagonia showed an initial admixture between Native American and European with no posterior pulses. The ancestry decay (top) compares the expected and observed tract length distributions based on the best-fit model, with the shaded area indicating the 68.7% confidence interval. The points indicate the observed values. The migration model (bottom of each graph) shows the change in admixture proportion over time. From left to right along the time scale in generations ago (GA).

### Supplementary Note 9: Comparative Analysis of Selection Signals in Patagonian Populations and Known Adaptive Loci from Northern Indigenous Groups

To investigate signatures of local adaptation in present-day Patagonian populations, we conducted genome-wide selection scans using both allele frequency-based statistics (PBSn1, (Yi et al., 2010; Crawford et al., 2017)) (see Figure 4). As part of this analysis, we evaluated whether known candidate genes previously associated with adaptation to cold climates and lipid-rich diets in Northern Native American populations (Clemente et al., 2014, 2014; Fumagalli et al., 2015) were also under selection in our dataset. Our current dataset was already masked for all non-Native American ancestries as previously described (see Supplementary Note 5).

We focused on two population subsets: (1) all Patagonian individuals (Supplementary Figure S10), and (2) maritime hunter-gatherer groups from western Patagonia (Laitec, Yámana, and Kawésqar; Supplementary Figure S11). Specifically, we tested for selection signals in well-characterized loci such as the FADS gene cluster (chromosome 11), LRP5 and CPT1A (chromosome 11), and THADA (chromosome 2), which have been implicated in lipid metabolism and thermoregulation in Arctic and sub-Arctic populations.

While no SNPs within these candidate genes surpassed the top 1% genome-wide significance threshold in our selection scans, we did observe clusters of SNPs near the FADS region with elevated PBSn1 and iHS scores. These suggest potential signals of moderate selection proximal to this functionally relevant locus, though not strong enough to meet genome-wide criteria for outlier detection in our current dataset.

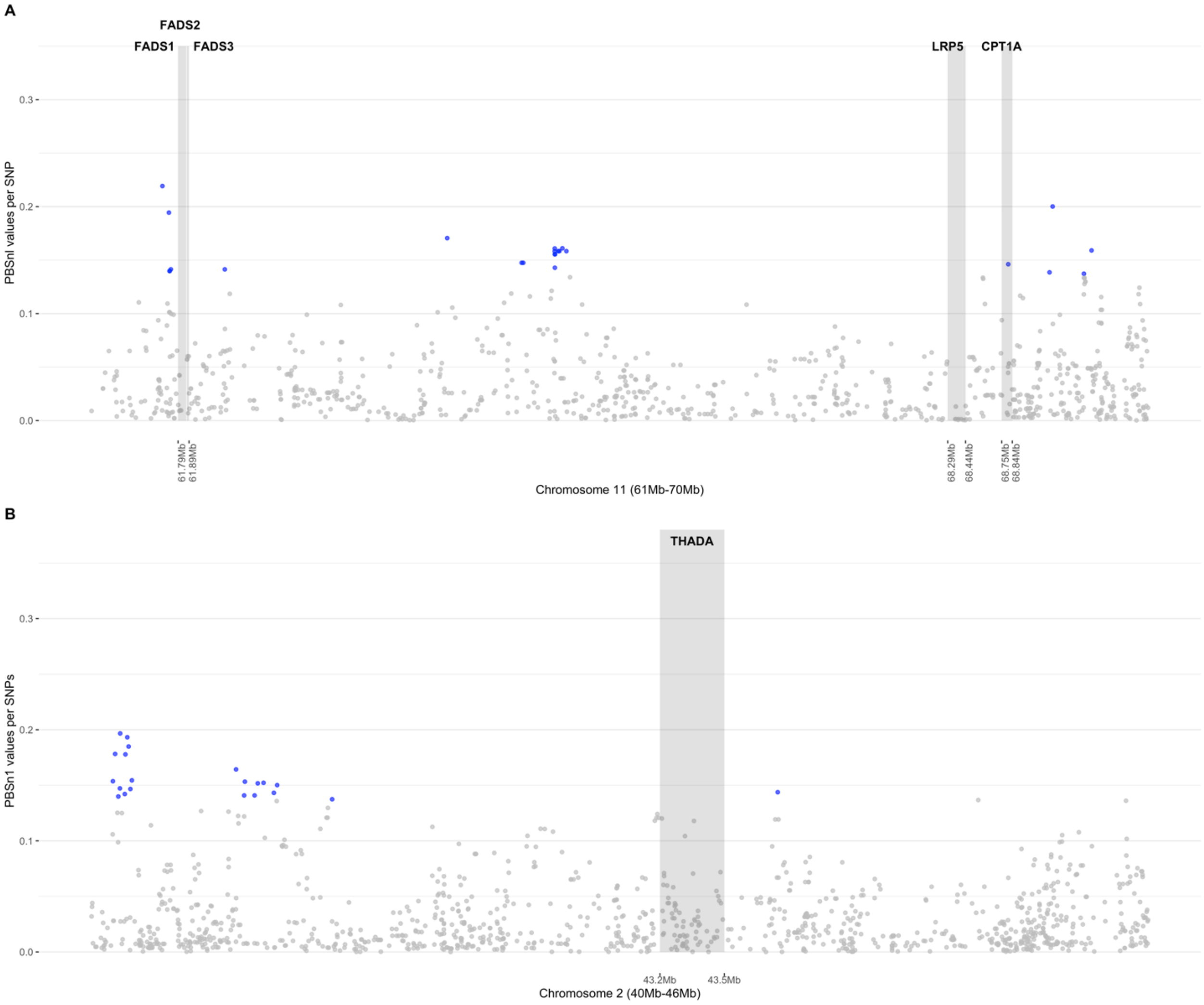


**Figure S9 | Population Branch Statistics (PBSn1) in Patagonian hunter-gatherers.**
Comparison of PBSn1 (Yi et al., 2010; Crawford et al., 2017) scores in Patagonian hunter-gatherers' individuals with respect to previous adaptive candidate genes. Each dot represents a PBSn1 value per SNP where the color denotes if it is above percentile 1% (blue) or not (gray). The genes related to the cold extreme and maritime diet in Siberia in chromosomes 11 (A) and 2 (B) are drawn in grey rectangles.

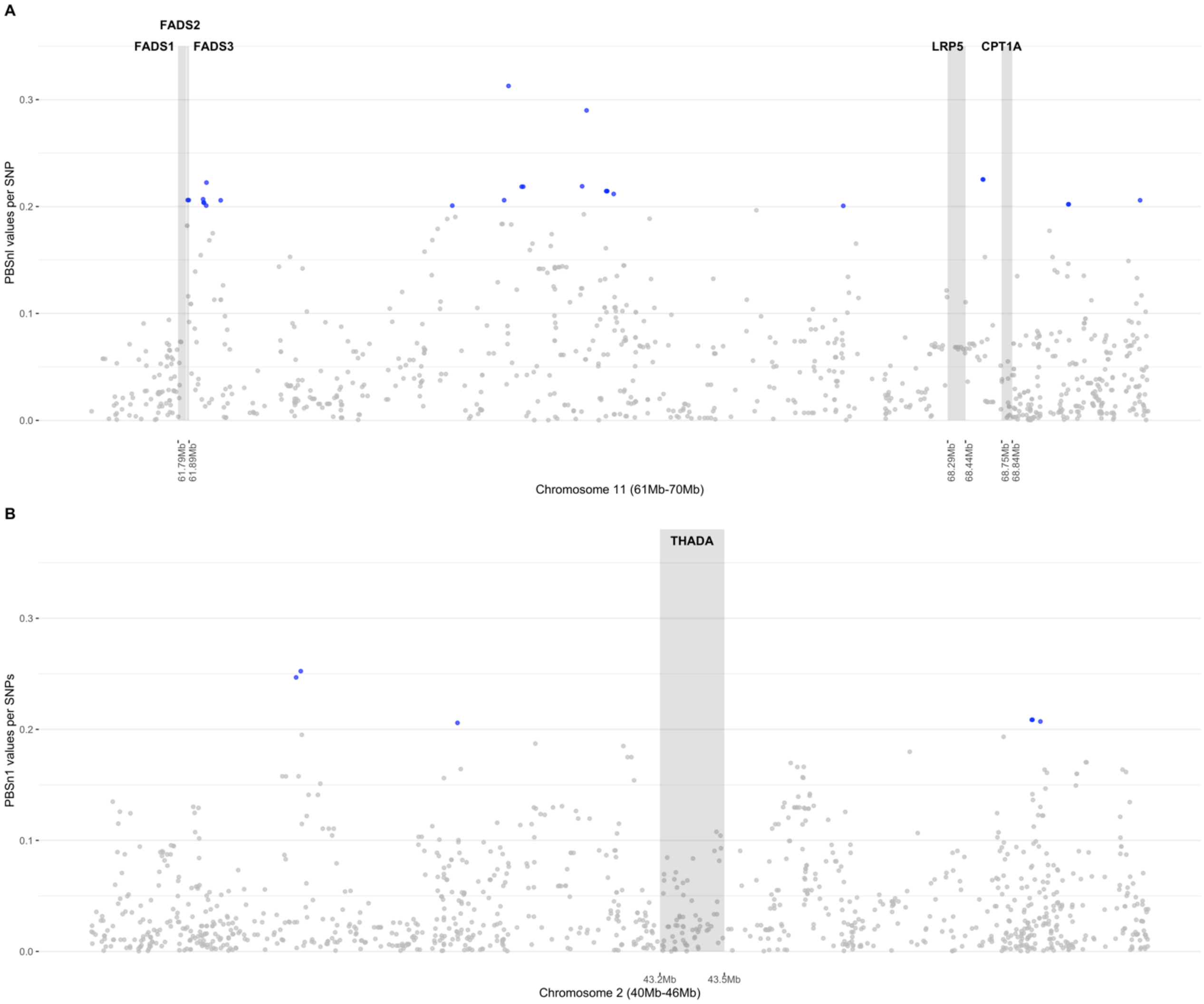


**Figure S10 | Population Branch Statistics (PBSn1) in maritime hunter-gatherers.** Comparison of PBSn1 (Yi et al., 2010; Crawford et al., 2017) scores in maritime hunter-gatherers' individuals from Patagonia with respect to previous adaptive candidate genes. Each dot represents a PBSn1 value per SNP where the color denotes if it is above percentile 1% (blue) or not (gray). The genes related to cold extreme and maritime diet in Siberia in chromosomes 11 (A) and 2 (B) are drawn in grey rectangles.

**Supplementary Note 10: Comparative Analysis of Genomic Selection Signals in Ancient and Present-Day Patagonian Populations**

To assess the temporal persistence of adaptive signals, we compared selection signatures in ancient individuals from southern Patagonia and Tierra del Fuego—sourced from the AADR database (Mallick et al., 2023)—with those identified in present-day Patagonian individuals. We focused on PBSn1-based selection scores and generated pairwise scatterplots to visualize patterns of overlap between the two datasets, stratified by Patagonian hunter-gatherer populations and maritime hunter-gatherers (Supplementary Figures S12–S13).

Regions where both ancient and modern individuals exhibited PBSn1 values exceeding the top 1% threshold were interpreted as candidates for long-term selection continuity. Although several loci showed concordant elevated values across time, none of these regions overlapped with the top-scoring selection peaks in present-day individuals, suggesting either attenuation of historical selective pressures or demographic dilution of ancestral signals.

It is important to note that small sample sizes, particularly in ancient groups, and the high degree of missingness in the AADR-derived ancient genotypes may limit power to detect or replicate true adaptive regions. Nonetheless, many of the overlapping loci–albeit at moderate significance–were functionally linked to lipid metabolism and immune response, consistent with known adaptive pressures in both ancient and contemporary Patagonian environments.

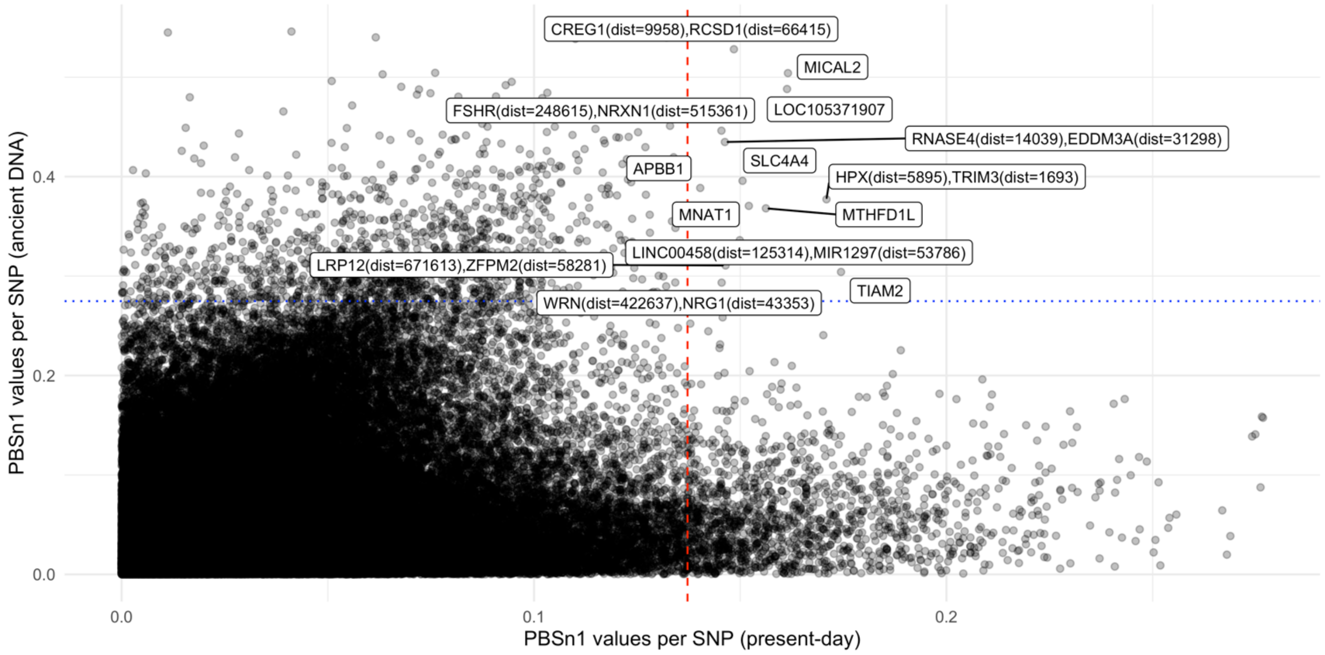


**Figure S11 | Scatterplot of PBSn1 SNP scores between ancient and present-day representative for Patagonian hunter-gatherers.**

Comparison of PBSn1 (Crawford et al., 2017; Yi et al., 2010) scores in Patagonian hunter-gatherers' descendants (x-axis) against the ancient samples from Patagonia and Tierra del Fuego (y-axis). Each dot represents a PBSn1 value per SNP in common from both datasets. All dots labeled with their genes fall within the top 1% distribution of the data.

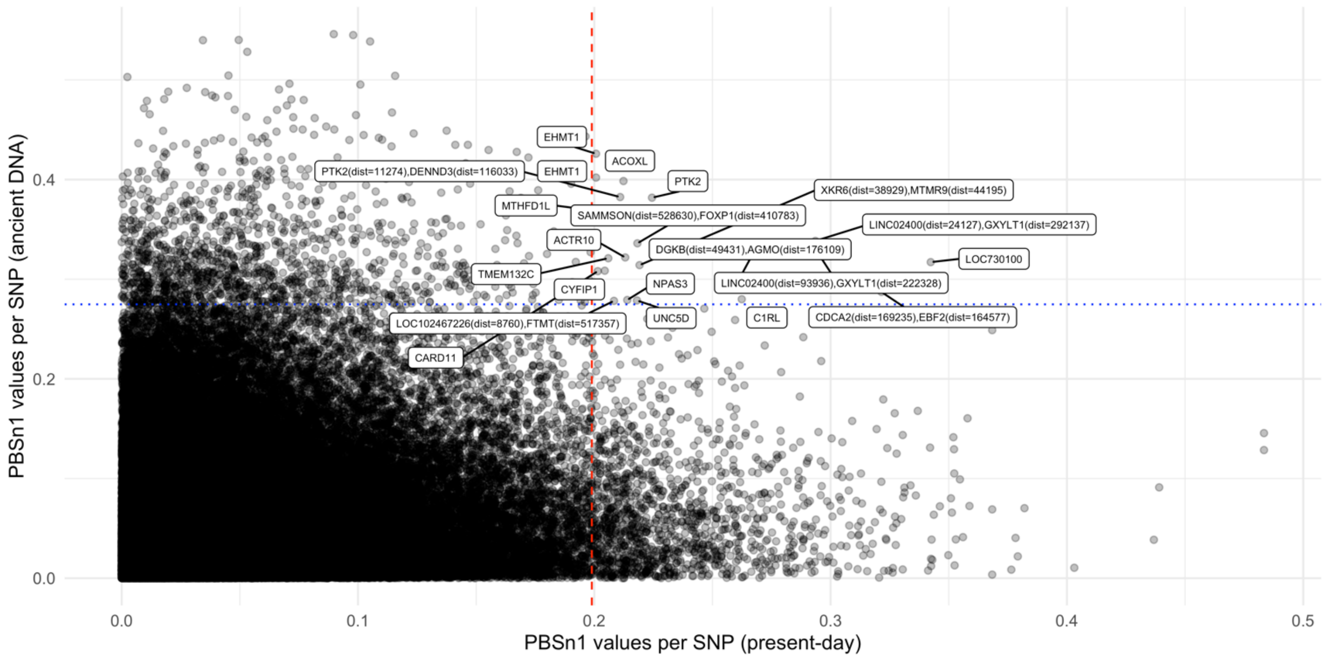


**Figure S12 | Scatterplot of PBSn1 SNP scores between ancient and present-day representatives for maritime hunter-gatherers.**

Scatterplot comparison of PBSn1 (Yi et al., 2010; Crawford et al., 2017) scores in maritime hunter-gatherers' descendants from Patagonia (x-axis) against the ancient samples from Patagonia and Tierra del Fuego (y-axis). Each dot represents a PBSn1 value per SNP in common from both datasets. All dots labeled with their genes fall within the top 1% distribution of the data.

### Supplementary Note 11: Population Branch Statistic (PBS)

To identify loci under population-specific positive selection, we applied the Population Branch Statistic (*PBSn1*), a widely used allele frequency-based method that quantifies excess allele frequency divergence in a focal population relative to a closely related reference population and a distantly related outgroup. PBS assumes that, under neutrality, allele frequency differences between populations follow expectations based on genetic drift; significant deviations from this model suggest population-specific adaptation (Yi et al., 2010).

PBSn1 computes branch-specific divergence by transforming pairwise Fst distances into drift time estimates, enabling detection of selection along a specific lineage while controlling for shared demographic history. Higher PBS values indicate loci where the focal population exhibits stronger divergence than expected under neutrality.

In Supplementary Figure S14, we present the PBSn1 framework in two demographic configurations: (A) for Patagonian hunter-gatherers, with the focal branch being Patagonian hunter-gatherers, the sister branch comprising South-Central Chilean populations (Pehuenche-Huilliche), and the Han as the outgroup; and (B) for maritime hunter-gatherers (Yámana, Kawésqar, Laitec), with the Argentine population as the sister branch and Han as the outgroup. This dual approach allowed us to explore environment-specific selection pressures, including those potentially linked to marine adaptation, subsistence changes, and post-glacial recolonization dynamics.

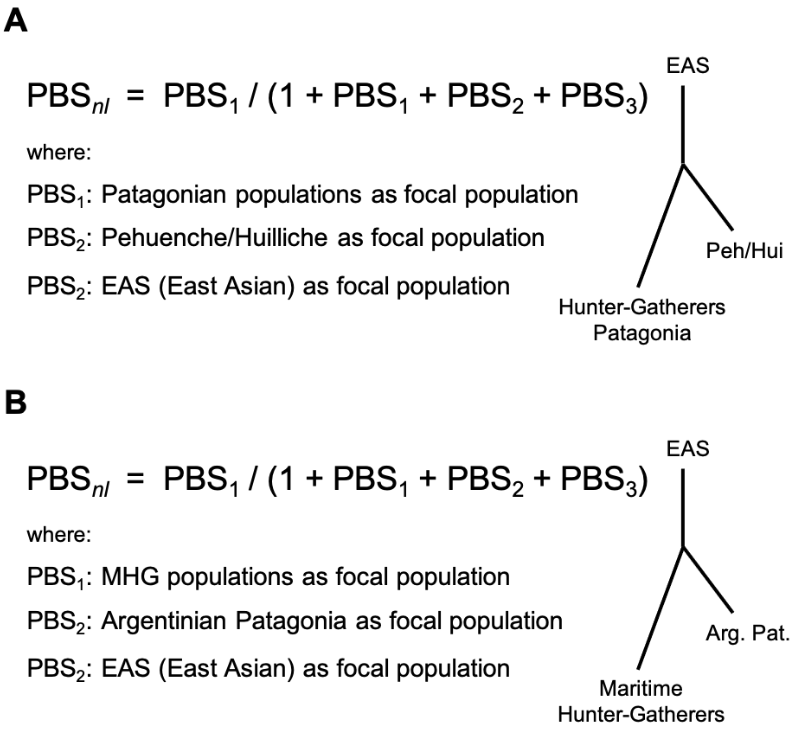


**Figure S13 | PBSnl scheme for Patagonian populations.**

In A is shown the first scenario for the entire Patagonia (focal), Pehuenche and Huilliche (Peh/Hui) as near group and Han Chinese (EAS) as outgroup. In B is shown the second scenario with maritime hunter-gatherers (MHG) as a focal group, Argentinian Patagonia (Pat. Arg) as a near group and Han Chinese as the outgroup.

FACULTAD MEDICINA
UNIVERSIDAD DE CHILE

Laboratorio ChileGenomico
Programa de Genética Humana, ICBM
Av. Independencia 1027
Santiago, Chile
http://genomed.med.uchile.cl

INSTITUTO DE CIENCIAS BIOMÉDICAS

### APPLICATION FOR ACCESS TO THE DATA INCLUDED IN THE PAPER:

Genomic evidence of human adaptation before and after European arrival in present-day Patagonian populations

Applications for access to these data can be submitted at any time. These will be considered on a rolling basis and we aim to provide a decision within 1 month of receipt.

Name of applicant and co-applicant(s), including affiliations and contact details.

Enter the Primary E-mail correspondence E-mail address you would like to use

PhD student applicants must include their supervisors as a co-applicant and provide their full contact details.

**Title of Project (in less than 30 words).**

Please provide a clear description of the project and its specific aims in no more than 750 words. This should include specific details of what you plan to do with the data and include key references.

**Research ethics**

Do you foresee any ethical issues, such as potential stigmatization of ethnic groups, arising as a result of your research? If so, how to you plan to address such issues?

**If this application is accepted:**

(a) I/we will not secondarily distribute the data to anyone
(b) I/we will not post it publicly
(c) I/we will make no attempt to connect the genetic data to personal identifiers for the samples
(d) I/we will not use the data for any commercial purposes.

Violation of this agreement is considered a serious breach of ethical conduct and may lead to legal action.

**Name:**

**Date:**

**Signature:**

---

UNIVERSIDAD DE CHILE USE ONLY:

Date request received:

Data request number:

Date data request approved:

Date data transmitted:

### References

,mn
